# Relationship of plasma biomarkers to digital cognitive tests in alzheimer’s disease

**DOI:** 10.1101/2023.08.21.23293786

**Authors:** Sofia Toniolo, Sijia Zhao, Anna Scholcz, Benazir Amein, Akke Ganse-Dumrath, Amanda J Heslegrave, Sian Thompson, Sanjay Manohar, Henrik Zetterberg, Masud Husain

**Affiliations:** Nuffield Department of Clinical Neurosciences, University of Oxford, Headley Way, Headington, OX3 9DU, Oxford, United Kingdom; Cognitive Disorders Clinic, JR Hospital, Headley Way, Headington, OX3 9DU, Oxford, United Kingdom; Department of Experimental Psychology, University of Oxford, Woodstock Road, OX2 6GG, Oxford, United Kingdom; UK Dementia Research Institute, UCL, Gower Street, WC1E 6BT, London, United Kingdom; Department of Neurodegenerative Disease, UCL Institute of Neurology, Queen Square, WC1E 6BT, London, United Kingdom; Institute of Neuroscience and Physiology, University of Gothenburg, Huvudbyggnad Vasaparken, Universitetsplatsen 1, 405 30, Gothenburg, Sweden; Clinical Neurochemistry Laboratory, Sahlgrenska University Hospital, Göteborgsvägen 31, 431 80, Mölndal, Sweden; Hong Kong Center for Neurodegenerative Diseases, 15/F Building 17W, 17 Science Park W Ave, Science Park, Hong Kong, China; Wisconsin Alzheimer’s Disease Research Center, University of Wisconsin School of Medicine and Public Health, University of Wisconsin-Madison, 600 Highland Ave, J5/1 Mezzanine, 53792, Madison, WI, USA

**Keywords:** Cognition, phospho tau, memory, dementia, online testing

## Abstract

**Introduction:** A major limitation in Alzheimer’s disease (AD) research is the lack of the ability to measure cognitive performance at scale - robustly, remotely, and frequently. Currently, there are no established online digital platforms validated against plasma biomarkers of AD.

**Methods:** We used a novel web-based platform that assessed different cognitive functions in AD patients (N=46) and elderly controls (N=53) who were also evaluated for plasma biomarkers (amyloid β42/40 ratio, pTau181, GFAP, NfL). Their cognitive performance was compared to a second, larger group of elderly controls (N=352).

**Results:** Patients with AD were significantly impaired across all digital cognitive tests, with performance correlating with plasma biomarker levels, particularly pTau181. The combination of pTau181 and the single best-performing digital test achieved high accuracy in group classification.

**Discussion:** These findings show how online testing can now be deployed in patients with AD to measure cognitive function effectively and related to blood biomarkers of the disease.

## 1 Background

The advent of new disease-modifying treatments for Alzheimer’s disease (AD) has highlighted the need for sensitive cognitive tests to stratify those who might benefit from early interventions [1,2]. Traditional face-to-face neuropsychological assessments can detect changes only several years after pathological accumulation of amyloid and tau, a factor that might have led to past clinical trial failures [3]. Digital cognitive metrics capture subtle signs of impairment that cannot be measured by standard clinical tests and might be particularly valuable in early phases of the disease when cognitive impairment is at subthreshold levels on current scales [4]. Screening for AD, recruitment and longitudinal follow-up in clinical trials could all be transformed if cognitive testing were to be conducted robustly, remotely, and frequently [5].

Recognized biomarkers for AD diagnosis are currently divided into three main categories according to the ATN (amyloid, tau and neurodegeneration) staging system [6]; amyloid markers (reduced Aβ42 or Aβ42/Aβ40 ratio in the cerebrospinal fluid (CSF), or positive amyloid PET), tau (increased phosphorylated tau (p-Tau 181) in the CSF or positive Tau PET), and neurodegeneration (atrophy on magnetic resonance imaging (MRI), positive 18F-fluorodeoxyglucose (FDG) PET and increased total tau (t-Tau) in the CSF).

However, work on plasma biomarkers for AD has advanced rapidly. Regarding amyloid, plasma Aβ42/40 ratio has been found to correlate well with its CSF levels and with amyloid positron emission tomography (PET) findings, and its reduction is associated with cognitive decline in cognitively unimpaired individuals, people with subjective cognitive impairment (SCI), and mild cognitive impairment (MCI) [6–10].

Plasma pTau181 has been emerging as an even more specific and sensitive biomarker for AD [10–12]. It correlates well with its levels in the CSF [10], and is associated with both amyloid and tau PET positivity [10,12]. It is elevated in amyloid-positive individuals, even in the pre symptomatic stage, while it is not increased in several clinical mimics of AD [10,13].

Plasma glial fibrillary acidic protein (GFAP), which is a marker of neuroinflammation and reflects astrocytosis, is also associated with amyloid deposition in healthy controls, SCI, MCI and AD dementia patients [14–17]. Some evidence suggests that it is better than Aβ42/40 ratio in predicting amyloid PET positivity [14,17]. However, raised GFAP levels are not specific to AD, and are also increased in many other neurological diseases [18]. Similarly to GFAP, another plasma biomarker that is altered across several neurological disorders is neurofilament light chain (NfL). High baseline levels of NfL, an index of the rate of axonal injury, are strongly linked to markers of neurodegeneration such as CSF total tau (t-tau), MRI atrophy and FDG-PET hypometabolism [19–21].

Cognitive performance may be the single most important factor to increase the diagnostic accuracy of plasma biomarkers in AD, compared to other measures including MRI imaging and ApoE status [22]. Although some studies have examined the relationship between plasma biomarkers and cognition, to our knowledge, the cognitive tests used were not digital online measures. For example, baseline levels and longitudinal increases in plasma pTau181 are associated with a decline on standard tests of cognition such as the mini mental state examination (MMSE) [11,16,23–26]. The increase of another pTau isoform, plasma pTau217, was found to correlate with worsening cognition on the MMSE and modified Preclinical Alzheimer’s Cognitive Composite (mPACC) [27] in cognitively unimpaired and MCI participants. High GFAP and NfL levels have also been linked to worse cognitive performance on standard tests of cognition [17,28,29], to a decline in cognition over time [19,21,30,31], and clinical conversion to AD [15]. However, these changes do not seem to be AD-specific [32].

A head-to-head study comparing different plasma biomarkers in cognitively unimpaired individuals with positive amyloid status [33] found that pTau217, pTau181, and GFAP were associated with cognitive decline (on the mPACC and MMSE), while pTau 231 and NfL were not. pTau217 was the strongest individual predictor of cognitive impairment. However, standard neuropsychological tests such as the mPACC and the MMSE still require a dedicated face-to-face appointment, which limits their use for large-scale population screening. While the associations with standard cognitive metrics might shed a light on the ability of specific biomarkers to capture overall global cognitive impairment using standard pen-and-paper tests, research is needed on whether this applies to digital tests as well. Currently, to the best of our knowledge, there are no published studies which have reported on the association between plasma biomarkers and performance on a panel of digital cognitive tests. Additionally, the presence of apathy and depression, particularly in patient populations, might result in lower engagement in online testing, but whether their presence affects digital tests performance remains unknown.

Here, we investigated whether plasma biomarkers (Aβ42/40 ratio, pTau181, GFAP, and NfL) are associated with several digital online cognitive metrics, measuring visual short-term memory, long-term memory, visuospatial copying, executive function, and processing speed in a cohort of patients with AD and two samples of elderly healthy controls, one of which also underwent blood collection for plasma biomarker measurement. While cognitive impairment has historically been a key feature of all previous diagnostic criteria for AD [7,8], according to the ATN criteria its presence is not necessary to support AD diagnosis. Conversely, AD as defined by the in vivo detection of the accumulation of amyloid and tau, can occur even in asymptomatic individuals. Here, in a real-world cohort of patients, we used the standard clinical criteria for diagnosis of probable AD [34] which do not require CSF or PET imaging evidence of ATN positivity, and examined the relationship of clinical classification to performance on our digital cognitive tests and plasma biomarkers of AD.

## 2 Methods

### 2.1 Ethics

The study was performed in accordance with the ethical standards as laid down in the 1964 Declaration of Helsinki and its later amendments. Ethical approval was granted by the University of Oxford ethics committee (IRAS ID: 248379, Ethics Approval Reference: 18/SC/0448). All participants gave written informed consent prior to the start of the study.

### 2.2 Participants

46 patients with AD and 53 elderly healthy controls (EHC) were recruited respectively from the Cognitive Disorders Clinic (AD) at the John Radcliffe Hospital in Oxford, United Kingdom, or open day events (EHC). Patients with AD had a progressive, multidomain, largely amnestic cognitive impairment and underwent MRI and FDG-PET imaging, the results of which were in keeping with a clinical diagnosis of AD (temporo-parietal atrophy and hypometabolism) [34], but did not have ATN confirmation prior to enrollment in the study. Elderly healthy controls were > 50 years old, had no psychiatric or neurological illness, were not on regular psychoactive drugs, and all scored above the cut-off for normality (88/100 total Addenbrooke’s Cognitive Examination-III (ACE) score), [35]. They also underwent brain MRI imaging, and only participants with a normal MRI scan, reviewed by two independent senior neurologists, were included in the study. The groups were not statistically different in age, gender, or education level (Table 1).

**Table 1.** Demographics, plasma biomarkers, standard cognitive metrics and questionnaire-derived apathy and depression scores. All metrics are reported in group mean and standard deviation. * indicates p-values below <0.001, n.s. means not significant. The effect size for group comparison is rank-biserial correlation coefficient. ACE: The Addenbrooke’s Cognitive Examination-III. AMI: Apathy Motivation Index. GDS: Geriatric Depression Scale.

| Dimensions | Metrics | AD (n= 46) | EHC 1 (n= 53) | EHC 2 (n=352) | AD vs EHC 1 |  |
| --- | --- | --- | --- | --- | --- | --- |
|  |  |  |  |  | p value | Rank-Biserial Correlation |
| Demographics | Age | 68.3 (10.2) | 68.6 (7.0) | 59.9 (8.5) | n.s. |  |
|  | Gender (M/F) | 23/23 | 24/29 | 177/175 | n.s. |  |
|  | Education | 14.5 (3.5) | 15.8 (3.1) | 15.0 (2.1) | n.s. |  |
| Plasma biomarkers | pTau181 (pg/ml) | 5.4 (3.4) | 2.6 (1.3) | not applicable | * < 0.001 | -0.70 |
|  | GFAP (pg/ml) | 224.7 (118) | 111.8 (57.9) |  | * < 0.001 | -0.69 |
|  | NfL (pg/ml) | 29.7 (19) | 17.1 (11) |  | * < 0.001 | -0.58 |
|  | Aβ<sub>42/40</sub> ratio | 0.059 (0.01) | 0.067 (0.01) |  | * < 0.001 | 0.53 |
|  | Aβ<sub>42</sub> (pg/ml) | 6.6 (1.5) | 7.1 (1.5) |  | 0.06 | 0.22 |
|  | Aβ<sub>40</sub> (pg/ml) | 115.2 (25.7) | 108.4 (19.9) |  | n.s. | -0.13 |
| ACE | Total score | 63.5 (20.0) | 97.4 (2.0) | not applicable | * < 0.001 | 0.93 |
|  | Attention | 12.0 (4.4) | 17.2 (0.3) |  | * < 0.001 | 0.88 |
|  | Memory | 11.7 (6.1) | 25.0 (1.1) |  | * < 0.001 | 0.95 |
|  | Fluency | 7.4 (3.6) | 13.2 (0.9) |  | * < 0.001 | 0.89 |
|  | Language | 21.0 (4.9) | 25.6 (0.7) |  | * < 0.001 | 0.78 |
|  | Visuospatial | 11.9 (4.2) | 15.8 (0.3) |  | * < 0.001 | 0.79 |
| Questionnaires | AMI | 1.5 (0.3) | 1.2 (0.4) | 1.5 (0.4) | 0.002 | -0.40 |
|  | GDS | 6.9 (1.9) | 4.9 (1.1) | 5.7 (2.0) | * < 0.001 | -0.64 |

Participants underwent blood collection, face-to-face standard cognitive, and online remote digital cognitive testing, and self-reported motivation and mood indices were collected (see Figure 1 for study flow).

**Figure 1.**
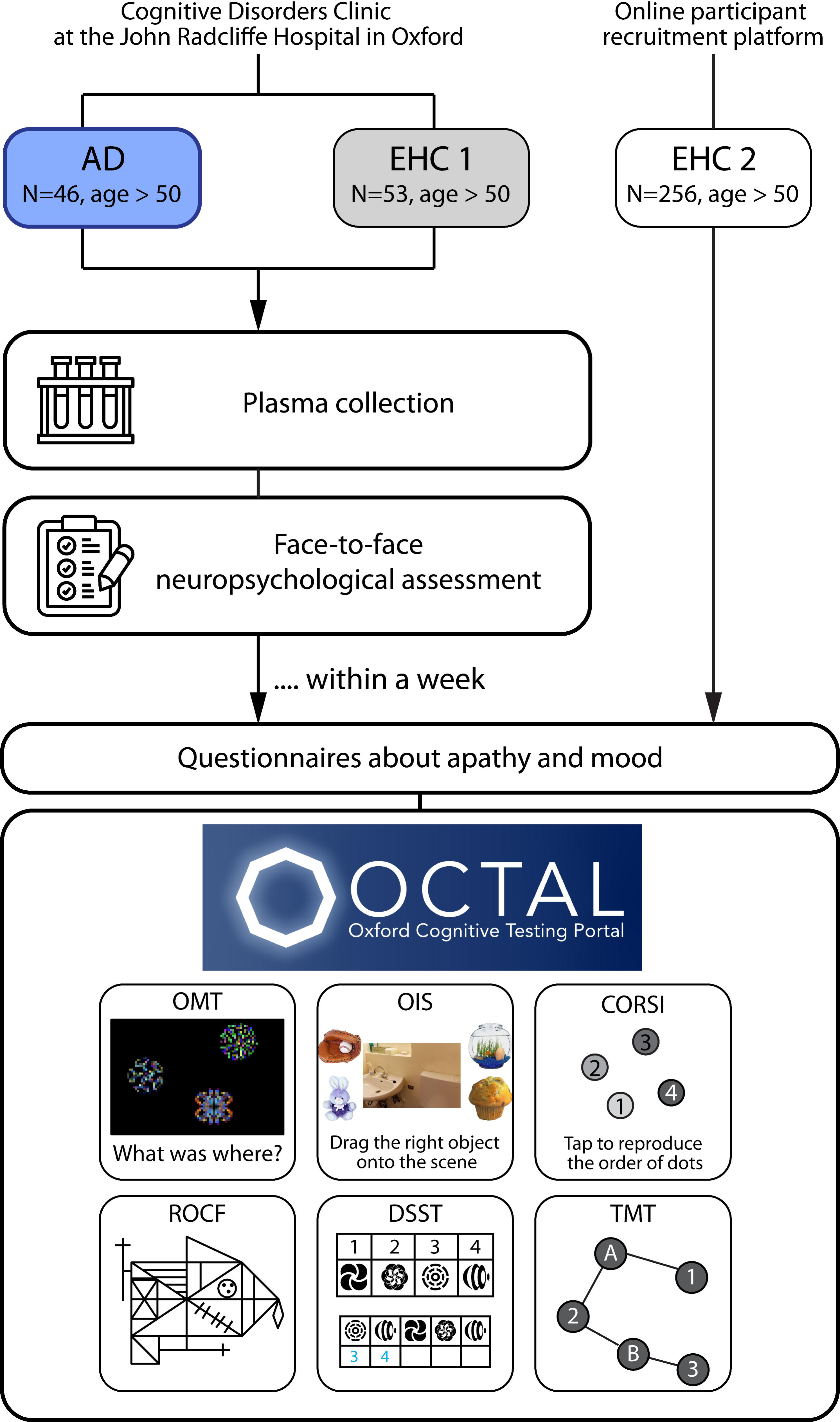
Study schematic. AD = Alzheimer’s disease. EHC = Elderly Healthy Control.

Since human behavior sampled for convenience only across university populations may be WEIRD (Western, Educated, Industrialized, Rich and Democratic) [36], the healthy controls recruited through the University (EHC1) may not necessarily be representative of the general population. Therefore, we recruited 356 healthy participants above 50 years old online through the Prolific participant recruitment platform (prolific.co) as a second normative group (group EHC 2, see Table 1 for basic demographics). All participants were blinded with respect to the aim of this study which was advertised as “a brain game” testing how well people could perform. They had normal or corrected-to-normal vision acuity and no color blindness. All were residents of the UK, had English as their first language and self-reported to be neurologically healthy. Four participants from this group were excluded due to failing multiple attention checks (see 2.7 Attention checks), leaving this group 352 valid participants. This EHC2 group was well-representative of ethnic groups and subjective socioeconomic status of the general public of the UK (see Supplementary Methods 1 for more details). To account for the effect of age on cognitive metrics, the cognitive performance of all participants was transformed into z-score based on EHC 2 group (see details below “Normalisation for digital cognitive metrics”; Table 2 and Supplementary Table 1).

**Table 2.** Digital cognitive metrics from OCTAL. All metrics were normalised by age-matched normative data (i.e., participants who are EHC 2). All metrics are reported in group mean and standard deviation. * indicates p-values below <0.001. The effect size for group comparison is rank-biserial correlation coefficient. See supplementary table 1 for the extended table, including all cognitive metrics and the rank-biserial correlation coefficients for group comparison with EHC 2. OMT: Oxford Memory Test; OIS: Object-in-Scene task; ROCF: Rey-Osterrieth Complex Figure; CORSI: Freestyle Corsi tapping task; DSST: Digital Symbol Substitution Task; TMT: Trail-Making task.

| Digital Cognitive Tasks | Metrics | AD (n=46) | EHC 1 (n= 53) | EHC 2 (n=352) | AD vs EHC 1 | AD vs EHC 2 | EHC 1 vs EHC 2 |
| --- | --- | --- | --- | --- | --- | --- | --- |
| OMT | Identification Accuracy (z) | -1.3 (1.9) | 0.6 (1.0) | -0.2 (1.4) | r = 0.59, p <0.0001 | r = 0.38, p = 0.0027 | r = -0.37, p <0.0001 |
|  | Location Error (z) | 4.4 (3.4) | 0.9 (1.7) | 0.2 (1.4) | r = -0.64, p <0.0001 | r = -0.77, p <0.0001 | r = -0.33, p = 0.0002 |
|  | Target Detection Rate (z) | -1.3 (1.6) | 0.7 (0.9) | -0.1 (1.2) | r = 0.71, p <0.0001 | r = 0.47, p = 0.0002 | r = -0.46, p <0.0001 |
|  | Misbinding Error Rate(z) | 36.9 (11.8) | 21.7 (7.9) | 28.1 (10.2) | r = -0.68, p <0.0001 | r = -0.47, p = 0.0001 | r = 0.42, p <0.0001 |
| **OIS** | Object Identification Accuracy<br>- Delayed Recall (z) | -2.5 (1.3) | 0.0<br>(1.2) | -0.1<br>(1.1) | $r = 0.82, p < 0.0001$ | $r = 0.82, p < 0.0001$ | $r = -0.07, n.s.$ |
| | Location Error - Delayed Recall<br>(z) | 4.0 (3.5) | 0.2<br>(1.0) | 0.3 (1.5) | $r = -0.70, p < 0.0001$ | $r = -0.68, p < 0.0001$ | $r = -0.05, n.s.$ |
| **ROCF** | Copy Raw Score (%) | 78.5<br>(30.2) | 98.3<br>(3.1) | 97.3<br>(4.9) | $r = 0.66, p < 0.0001$ | $r = 0.57, p < 0.0001$ | $r = -0.11, n.s.$ |
| | Copy Score (z) | -5.5 (8.8) | 0.1<br>(0.9) | -0.3<br>(1.6) | $r = 0.67, p < 0.0001$ | $r = 0.57, p < 0.0001$ | $r = -0.06, n.s.$ |
| | Immediate Recall Raw Score<br>(%) | 43.1<br>(22.8) | 89.9<br>(11.8) | 86.1<br>(13.2) | $r = 0.92, p < 0.0001$ | $r = 0.88, p < 0.0001$ | $r = -0.17, n.s.$ |
| | Immediate Recall Score (z) | -4.4 (2.7) | 0.3 (1.0) | -0.2<br>(1.3) | $r = 0.89, p < 0.0001$ | $r = 0.83, p < 0.0001$ | $r = -0.29, p = 0.0007$ |
| **CORSI** | Mean Location Error (z) | 2.0 (1.7) | -0.2<br>(0.8) | 0.3 (1.5) | $r = -0.77, p < 0.0001$ | $r = -0.64, p < 0.0001$ | $r = 0.09, n.s.$ |
| **DSST** | Number of Correct Responses<br>(z) | -2.3 (1.1) | -0.2<br>(0.9) | -0.0<br>(1.2) | $r = 0.86, p < 0.0001$ | $r = 0.87, p < 0.0001$ | $r = 0.10, n.s.$ |
| **TMT** | Average Completion Time for<br>TMT-A (z) | 4.3 (5.3) | 0.5 (1.8) | 0.2 (1.3) | $r = -0.79, p < 0.0001$ | $r = -0.82, p < 0.0001$ | $r = -0.12, n.s.$ |

### 2.3 Measurements of plasma biomarkers: Aβ42, Aβ40, pTau181, NfL and GFAP

Four plasma biomarkers were assayed:

● Amyloid pathology (‘A’): Aβ42, Aβ40 and the Aβ42/40 ratio, which is a better measure of amyloid pathology than Aβ42 alone [6,37].
● Tau pathology (‘T’): pTau181, which is a specific and sensitive marker of tau pathology in the blood and is highly predictive of tau PET positivity [11].
● Neurodegeneration (‘N’): NfL, the most commonly used blood-based biomarker reflecting the rate of neurodegeneration occurring in the brain [38].
● Astrocytosis: GFAP, an established marker of astrocytosis and synaptic plasticity [16].

### Plasma biomarker analysis

Blood was collected in 6 ethylenediaminetetraacetic acid (EDTA) tubes (10 ml each), and centrifuged (1800 g, RT, 10 minutes). The EDTA tubes were filled completely and gently inverted after collection to avoid coagulation. After centrifugation, plasma from all 6 tubes was transferred into one 50-mL polypropylene tube, mixed, aliquoted into 0.5 mL polypropylene tubes (Fluid X, Tri-coded Tube, Azenta Life Sciences), and stored at 4°C, until (< 8 hours) it was transferred into a -80°C freezer. The time between blood collection and centrifugation was < 30 minutes. Transfer time between 4°C and -80°C storage was < 20 minutes, and the samples were kept refrigerated during transport. All cryovials were anonymized, and the unique cryovial code was logged into a secure database, linked to the participant’s anonymous code and visit number.

Samples were shipped in dry ice to the Biomarker Factory / Fluid Biomarker Laboratory, UK Dementia Research Institute at University College London (UCL), London. The Dementia Research Institute (DRI) laboratory staff carried out the analyses. Plasma Aβ40, Aβ42, GFAP, and NfL were measured by Single molecule array (Simoa) technology using the Neurology 4-plexE assay on an HD-X analyser (Quanterix), according to manufacturer’s instructions. Plasma pTau181 was also measured by Simoa using the pTau-181 Advantage assay on an HD-X analyser (Quanterix). Further information regarding the analysis pipeline can be found in supplementary materials, in the ‘Supplementary Methods 2 section’.

### 2.4 Face-to-face neuropsychological screening

All participants completed the ACE in person at the time of the visit as a standard clinical screening test of cognition. ACE scores < 88/100 are considered to be abnormal, and all healthy controls scoring below that threshold were excluded from this study. However, patients with AD were not recruited based on a fixed threshold on standard cognitive testing but rather took part in the study according to the criteria outlined in paragraph 2.2, following clinical and radiological principles.

### 2.5 Digital cognitive test battery: Oxford Cognition Online Platform

Participants also completed a sequence of computerised cognitive tasks from OCTAL (Oxford Cognitive Testing Portal, available at https://octalportal.com) (Figure 2). The tasks include Oxford Memory Test (OMT), Object-in-Scene Memory Task (OIS), drag-and-drop version of Rey-Osterrieth Complex Figure (ROCF), Trail Making Test (TMT), Digit Symbol Substitution Test (DSST) and Freestyle Corsi Block Task (CORSI) (Figure 2). They measure distinct aspects of human cognition, various forms of memory, attention, and executive functions. They were adapted from established behavioural paradigms or neuropsychological tests, whilst being robust against the type of device that a person is tested on. These six tasks can be tried at https://octalportal.com. The tasks were conceived and designed by S.Z. and M.H.. Most of the tasks were built using the PsychoPy Builder (PsychoJS, version 2022.2.4) with custom-written codes in Javascript, with one exception: the Rey–Osterrieth Complex Figure (see details below), which was fully custom-written in HTML5 with JavaScript. All tasks were hosted on the server system Pavlovia.org.

**Figure 2.**
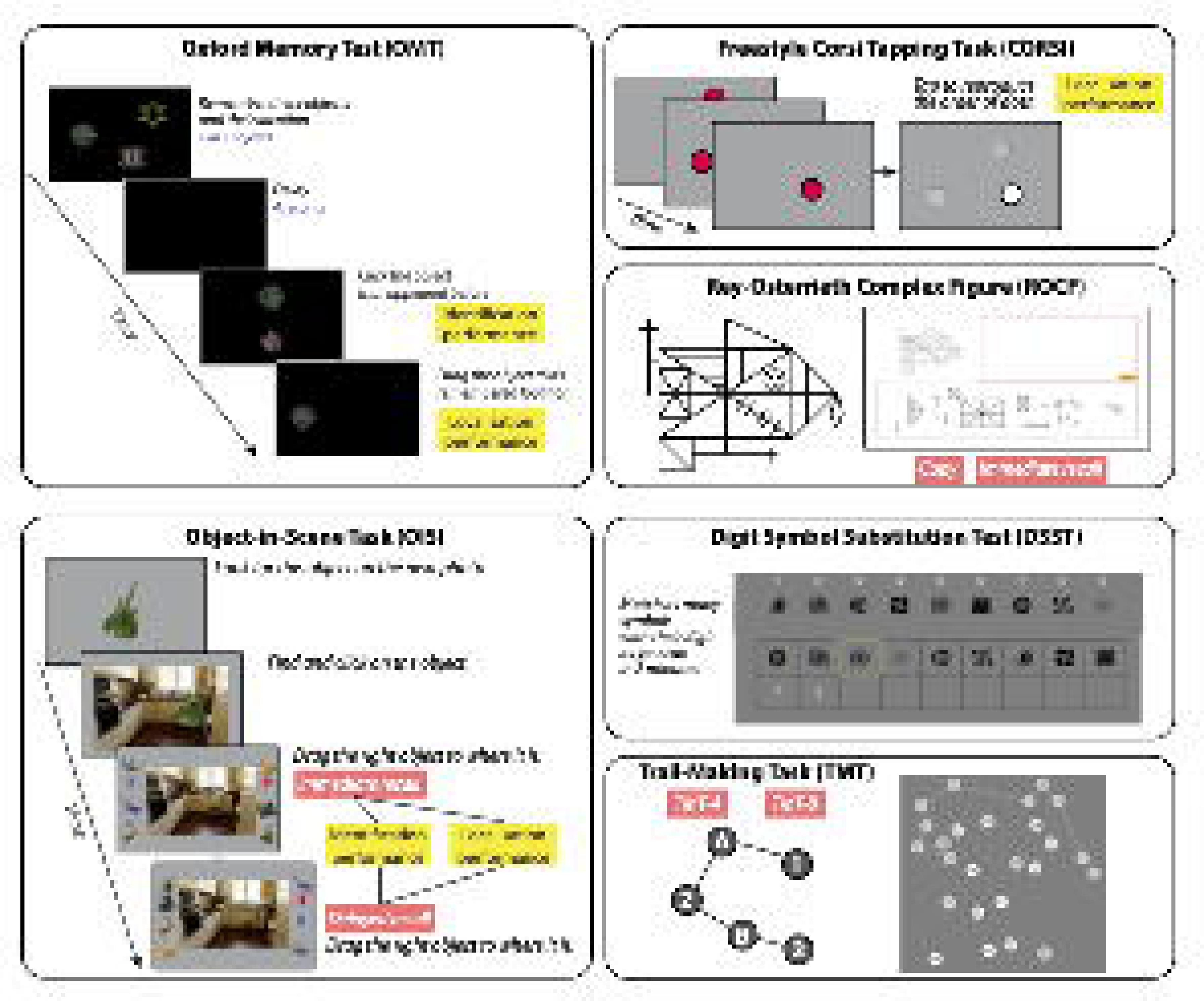
Experimental design of online digital cognitive tasks. Oxford Memory Test (OMT) is a “What was where?” visual short-term memory experiment where participants are presented with one or three fractal patterns positioned at various locations on-screen for 3 seconds. After a 4-second delay, participants identify the fractal pattern shown before and move it to its remembered location. Thus, the response reflects how precisely the memory was recalled. Additionally, the Object-in-Scene task (OIS) measures long-term memory. Participants are shown with a photo of an everyday scene and asked to remember a particular object placed in the picture. Another memory task is Freestyle Corsi Block Task (CORSI), which involves remembering a sequence of random locations on the screen. Participants are then free to click anywhere on-screen to indicate the remembered sequence of locations. Participants in the Rey-Osterrieth Complex Figure (ROCF) test their visuospatial abilities by dragging 13 elements given to copy a complex line graph on an empty canvas. Digit Symbol Substitution Test (DSST) requires participants to match symbols to digits according to a key located at the top of the page. More correct matches were made in under two minutes indicating faster processing speed. Trail Making Test (TMT) is also an executive function task. Participants are instructed to connect 25 circled numbers by clicking the circles in order as fast as possible. This task contains three trials of Task A (where the order is 1-2-3-4-5-6-…) and three trials of Task B (order 1-A-2-B-3-C-…). Full versions of all tasks can be tried online at https://octalportal.com.

A link with a unique patient and visit identifier was sent to the participants’ email address the same day as the in-person visit when blood was collected. Participants were required to use Chrome or Firefox browsers on a desktop, laptop or tablet with any operating system. They were encouraged to complete the online tests within a week maximum. After two weeks, the completion of the online tests was reviewed, and participants who did not complete the tasks within that time frame were prompted via email to do so. Tests completed more than 3 weeks after the blood samples were discarded.

#### 2.5.1 Oxford Memory Task (OMT)

OMT is the “What was where?” visual short-term memory experiment, which has been described in previous publications [4,39,40]. In this remote online version, participants were presented with one or three fractal patterns positioned at various locations on-screen for 3 seconds (Figure 2). Then, a 4-second delay was accompanied by a black screen. Subsequently, one of these fractal patterns was shown alongside a foil pattern. The two patterns were shown along the vertical meridian with 4 cm separation, with the order of the target and foil randomised across trials. Participants must identify which pattern they just saw (identification performance) by clicking the target pattern and drag it to its proper location on the screen (localisation performance). The foil was not a totally novel pattern; rather, it was part of the general pool of fractal images presented across the experiment. But the exact colour and shape of the foil never showed up as one of the patterns to remember.

Each participant performed a practice block of 6 trials including 3 trials with 1 item followed by 3 trials with 3 items. This is followed by a main test block of 40 trials, including 20 trials of 1 item and 20 trials of 3 items. The order of trials was randomised online. No feedback was given during practice or main test blocks. Fractal stimuli were drawn from a library of 196 pictures of fractals (http://sprott.physics.wisc.edu/fractals.htm), including 49 different shapes and each shape containing 4 colour variations.

As participants did the task remotely with their own devices, to ensure that the size of stimuli was physically the same across different devices, a card calibration procedure, previously described and validated [41], was employed prior to certain tasks. Participants are instructed to place a bank card or card of comparable size on the screen, and adjust the slider until the size of the image of the card on the screen matches the size of the physical card. This allows us to estimate screen distance by calculating the display’s logical pixel density in pixels per centimetre. After successful calibration, the diameter of the fractal stimulus is 2 cm. A Matlab script (MathWorks, Inc.) was used to determine the fractals’ locations in a pseudorandom manner with a few constraints. In order to avoid crowding and create a clear zone around the items’ original locations, which is essential for the analysis of localization errors, fractals were never placed closer than 3 cm to one another.

Eight cognitive metrics were extracted from this task: Identification Accuracy (proportion of correct object identification), Location Error (distance between response and target), Identification Time (reaction time to identify target), Localisation Time (reaction time to place object), Target Detection (rate of detecting correct object and placed at target location, see Figure 2), Misbinding (rate of placing target at a non-target location), Guessing (rate of placing target randomly) and Imprecision (how precise spatially is the response).

#### 2.5.2 Object-in-Scene Memory Task (OIS)

This test provides measures of identification accuracy, precision of spatial localization and semantic accuracy in visual short-term and long-term memory (Figure 2). Participants were presented with a photo of an everyday scene and instructed to remember a particular object placed in the picture. To aid effective encoding, the participant was also asked to click on the displayed object. Subsequently, 20 different objects were presented, and the participant was asked to choose the correct object and place it in the remembered location in the scene. To ensure that they were not simply remembering the name of the object, the object pool contained a foil that matches the target’s category (e.g., two guitars of different colour and shape). After 5 different object and scene pairs, participants were asked to reproduce the object-scene associations probed (delayed recall, after 3-4 minutes). There were a total of 20 trials divided into four blocks, and the order of the pairs was randomised.

Three metrics were extracted from the task for both immediate and delayed recall stages: object identification accuracy (proportion of trials in which participants correctly identified the original object; chance level = 5%), semantic identification accuracy (proportion of objects correctly recalled as belonging to the same semantic category as the target; chancel level = 10%), location error (the distance from the original target item location to the centre of participant’s response location; centimetre as unit).

#### 2.5.3 Rey-Osterrieth Complex Figure (ROCF)

This task is a digitised version of the traditional pen-and-paper test [42], which is an established measure of visuospatial abilities (Figure 2). The original ROCF task requires the participant to draw a complex line-drawing freehand, first by replicating an existing figure (copy), and then again from memory (immediate recall). Our digitised version does not require hand drawing. Instead, the figure is split into 13 independent elements, and participants are required to drag each element into an empty canvas to copy the figure. Each test was automatically scored using an offline MATLAB-based algorithm. In contrast to the discrete score used in the pen-and-paper version, the score of our digital version provides a continuous measure of precision. The middle large rectangle is selected as the anchor point as a reference element. If the element is not placed (not present on the canvas), there will be no score; otherwise, the distance from the large rectangle is computed. As a measure of imprecision for each element, the absolute difference between the ideal distance and the actual distance is then calculated. The absolute error is then scaled using a logarithmic function: if the element is placed relatively correctly, the difference in the distance from the big rectangle is computed; if the element is placed too far, the score is zero. The normalised absolute difference is then subtracted from 1 to calculate the score for this element. The sum of all element scores is 13, but the results are scaled to a percentage to match the original 36 element picture. Our version’s scoring is consistent with the pen-and-paper scoring guide, as the participant receives one point for correctly positioning the element and no score if the element is placed incorrectly or not at all on the canvas. This task and scoring have been validated with the in-person traditional 36 items pen-and-paper test and manual scoring with standard scoring guide in healthy participants before the start of the study. The percentages obtained at the copy and immediate recall – ROCF copy and recall scores – were used as metrics of interest.

#### 2.5.4 Digit Symbol Substitution Test (DSST)

DSST provides a measure of processing speed. In this digitised version, participants were required to match symbols to digits according to a key located at the top of the page (Figure 2). The key consisted of 9 symbols next to the digits 1–9. At the bottom of the screen, there was a row of 9 randomised symbols. Participants reported the digit that corresponded to each symbol by clicking on the correct digit. The row was refreshed once all 9 were answered. Participants were allowed two minutes to answer as many as they could and the number of correct matches within the allowed time (DSST - correct responses).

#### 2.5.5 Trail Making Test (TMT)

TMT is a standard test of processing speed and executive functions [43,44]. In this online version (Figure 2), 25 circled numbers are presented on-screen, and participants are instructed to connect them by clicking the circles in order as fast as possible. It contains three trials of Task A (where the order is 1-2-3-4-5-6-…) and three trials of Task B (order 1-A-2-B-3-C-…). Each participant sees six different trail maps randomly chosen from a pool of 100 pre-made maps, generated using a “divide-and-combine” approach [45]. The task also included a control condition of four trials to assess basic motor speed, where participants are presented with two circles located at two opposite corners of the screen. One is labelled with 1 and the other with 2, and participants were instructed to connect 1 with 2 as quickly as possible. The average of the reaction times of the TMT was used as a variable of interest.

#### 2.5.6 Freestyle Corsi Block Task (CORSI)

This task is a modification of the Corsi Block Tapping Task [46], which is a standard measure of visual short-term memory. In the original version, participants were presented with a set of nine identical wooden blocks positioned on a board. Subjects were required to point at the blocks in the order they were presented. They started with sequences of smaller blocks, and the number of blocks increased during the test. In the most computerised version of the task, the participant is shown several identical blocks that are in fixed locations spread across the screen [47]. Blocks then light up in sequence and the participant must remember which blocks lit up and in what order. In this digital version, the blocks’ locations were not fixed (‘Freestyle Corsi Tapping Task’). In an n-location trial, a 1-cm-wide red dot appeared at a random location on the screen, disappeared after 1 second, and reappeared at another random location on the screen (for n >1), and this process is repeated n times up to a sequence of 5 items. Once the sequence has finished, after a 1 second break the participant could freely click anywhere on-screen to indicate where each dot appeared in sequence. The location error was calculated as the average distance between the response and the target location. The task was divided into five blocks, each block having five trials of a n-location sequence (i.e., 5 blocks of 1-item, 5 blocks of 2 items, up to 5 blocks of 5 items). The average of the reaction times of the 5 conditions was chosen as a variable of interest.

### 2.6 Questionnaire-derived motivation and mood metrics

All participants also completed two questionnaires which were hosted on Qualtrics: (1) Apathy Motivation Index (AMI), an 18-item questionnaire, sub-divided into three subscales of apathy: emotional, behavioral and social apathy [48] and (2) Geriatric Depression Scale (GDS), short form, which includes 15 questions. It is a screening tool designed to assess depressive symptoms in elderly people. A total score greater than 5 indicates probable depression [49].

### 2.7 Attention checks

Attention checks were incorporated throughout the study: (1) In OIS, participants were required to adhere to the instruction of dragging an object onto the scene immediately after viewing the object and the photo. A low-effort response was defined as object identification accuracy lower than 20% (10% was the chance level for correct object and 20% was the chance level for correct semantic category). No participant failed this check. (2) In OMT, a failure of attention check was defined as an unreasonably short localisation time (cut-off at 0.2 seconds). Eight EHC2 participants failed this check. (3) In DSST, if the correct rate was below 20%, it meant the participants did not follow the instruction to match the digit with the symbol based on the key provided (the chance level for pure guessing would be 11%). Four EHC2 participants failed this attention check. (4) In DSST, if the participant stayed idle for more than 60 seconds in total, it was also considered to be a failure of attention check. One of the four participants who failed the last attention check also failed this check. (5) Healthy controls should be able to achieve a full score on ROCF copy. If a healthy control scored lower than 50%, their copy and recall would be excluded. All participants passed this check. (6) Each of the two questionnaires had a validation question “This is a validation question. Please choose ‘Completely untrue’.” No one failed this attention check. While no participant failed all six attention checks, four individuals were excluded after failing more than three of them. The performance of the tasks where attention checks failed was discarded.

### 2.8 Statistical analysis

For analysis and data visualisation purposes MATLAB (version R2023a), R studio (version 12.0), JASP (version 0.16.4) and SPSS (version 29.0) were used. Demographics, cognitive tests and plasma biomarkers levels were compared using a Mann-Whitney U test for continuous variables, while χ^2^ test was used for categorical variables such as gender. P-values were two-tailed with statistical significance set at p<0.05 for all analyses. Rank-biserial correlation was used for effect size estimation. If data from multiple visits was available, averaged values per participant across visits were used.

#### 2.8.1 Age-adjusted digital cognitive measures

Z-score (i.e. number of standard deviations from the mean of the normative population in the similar age (± 3 yrs)) was computed for each variable and each subject, based on a normative population of 352 online participants above 50 years old (EHC 2, see Table 1 for demographics). On average, each individual’s performance was adjusted with 55.8 (SD 14.7, min 30, max 81) participants from the EHC2 group.

#### 2.8.2 Correlation between digital cognitive metrics and plasma biomarkers

Plasma biomarkers’ log10 transformed values and Z-scores of digital cognitive measures were used for correlation and linear regression analyses. Correlations between digital cognitive metrics and plasma biomarkers were assessed with Spearman’s rank test, using age, sex, and education as covariates. The Benjamini–Hochberg method, which controls the False Discovery Rate (FDR), was used to correct for multiple comparisons.

#### 2.8.3 Machine learning for group classification

Additionally, machine learning was applied to predict group classification and plasma biomarkers levels, firstly using MATLAB-based algorithms for feature ranking, to estimate the absolute contribution of each variable. The *fscchi2* function in MATLAB (univariate feature ranking for classification using chi-square tests) was used to predict group classification, while the *fsrftest* function (univariate feature ranking for regression using F-tests) was used to predict continuous variables, i.e., plasma biomarkers’ levels. Rank’s importance scores were then transformed into p-values by calculating the exponential of the negative scores. Secondly, we applied the R-based MuMIn package to test which combinations of biomarkers would best predict group classification and plasma biomarkers levels. For predicting groups, we used logistic regression, while for predicting pTau181 level and Aβ42/40 ratio linear regression was deployed. The MuMIn package then uses the dredge function to achieve model selection, with the best performing model having the lowest corrected Akaike information criterion (AIC). ROC curves and AUCs were then computed for the model of interest. The pROC package in R with De Long’s test was used to compare model performance in direct comparisons between two ROC curves.

## 3 Results

### 3.1 Participants tests and plasma biomarkers overview

Patients with AD performed significantly worse in all digital cognitive metrics with high effect sizes (rank-biserial correlation, or rrb > 0.5) compared to matched controls (EHC1), see Figure 3 for distribution comparison for key cognitive metrics, and Supplementary Figures 1 and 2 for all metrics and online normative data. Patients with AD showed a large deficit in executive functions, indexed by trail making test (TMT) and digit symbol substitution (DSST), as indicated by on average 8.5 standard deviation (SD) below expectation. They also were 2.0-7.5 SDs below expectation in both identification and localisation of recalling remembered items in short-term memory (OMT and Freestyle Corsi Block Task (CORSI)) and long-term memory (Object-in-Scene Memory Task (OIS) delayed recall). Noticeably, patients with AD were particularly impaired at memory recall (OIS and Rey Osterrieth Complex Figure (ROCF)-recall). For example, both EHC groups could normally recall >90% of objects correctly with a very precise spatial memory (1 cm location error); in contrast, although AD remembered the object’s semantic category (for example, it was a guitar), they could only recall 72.9% of the objects (but which guitar?) accurately with an average location error of 7.5 cm away from the centre of the object (which is 2 cm wide). Similarly, EHCs recalled 80% of ROCF at immediate recall, but patients with AD on average scored less than 50% (5.2 SD below expectation).

**Figure 3.**
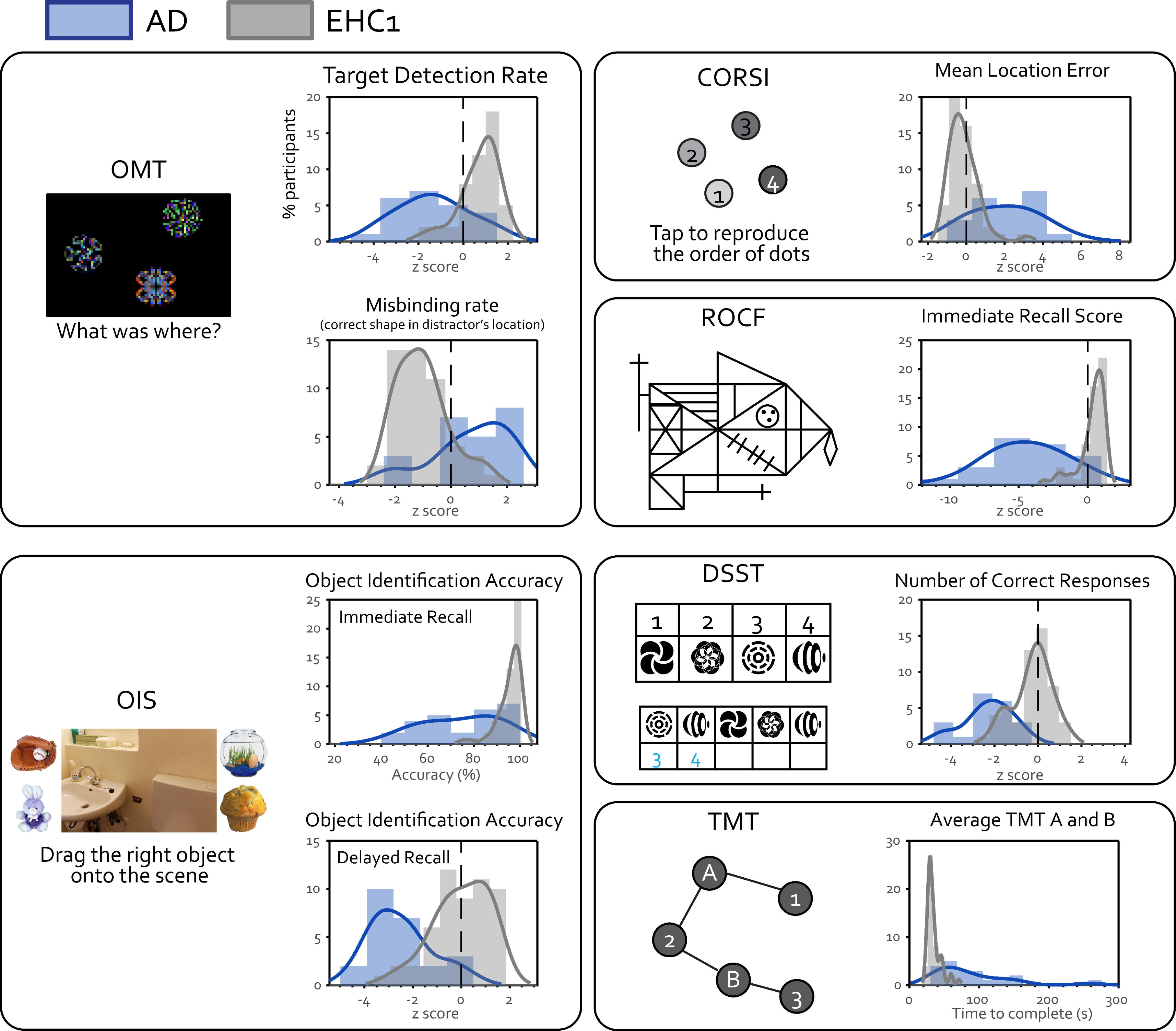
Digital cognitive metrics. In all online tasks, patients with AD (plotted in blue) performed significantly worse and had higher variability compared to age-matched healthy controls (EHC1, plotted in grey). On the X-axis, cognitive performance is shown as z-scores derived from age-matched online normative data, except TMT for which the raw completion time is shown. Y-axis indicates the percentage of participants. See supplementary materials for the distribution plots for all other cognitive metrics.

Compared to the online participants (EHC 2), EHC 1 performed slightly but significantly better in many cognitive metrics (see Supplementary Table 1). In our sample, this difference could not be explained by age, education level, or the testing environment (all completed remotely at home anonymously). EHC2 performed particularly worse in the Oxford Memory Task (OMT), where they made significantly more misbinding errors and were faster at localisation compared to the participants we tested locally. This group difference might be due to a speed-accuracy trade-off in EHC 2 group; in this online group, participants with shorter localisation time were associated with more misbinding errors (Pearson r = -0.22, p=0.003), while in contrast no correlation between speed and accuracy was found in EHC 1 (r=-0.07, p = 0.64).

Regarding plasma biomarkers, as expected, patients with AD had higher mean levels of pTau181, GFAP and NfL and lower Aβ42/40 ratio compared to cognitively unimpaired controls (Table 1).

ACE scores were significantly different between AD and EHC1, and had a high effect size in group comparisons, which was expected as it was the only test used for diagnosis (Table 1). Patients with AD were in general more apathetic and depressed compared to EHCs (Table 1). Across all participants, the apathy level overall strongly correlated with the level of depression (Pearson’s r = 0.46, p<0.001), but it did not significantly correlate with any of our online cognitive metrics (Supplementary Figure 3).

### 3.2 Relationships between plasma biomarkers and cognitive metrics

The relationships between all plasma biomarkers and digital cognitive metrics are visualised as a network plot in Figure 4a, in which the strength of the relationship is represented by the distance between the metrics. Among the four plasma biomarkers investigated in the present study, pTau181 was most strongly correlated with our digital cognitive metrics, which all clustered on the bottom right of the plot. In contrast, the Aβ42/40 ratio showed the weakest relationship with cognitive performance as well as with the other three plasma biomarkers.

**Figure 4.**
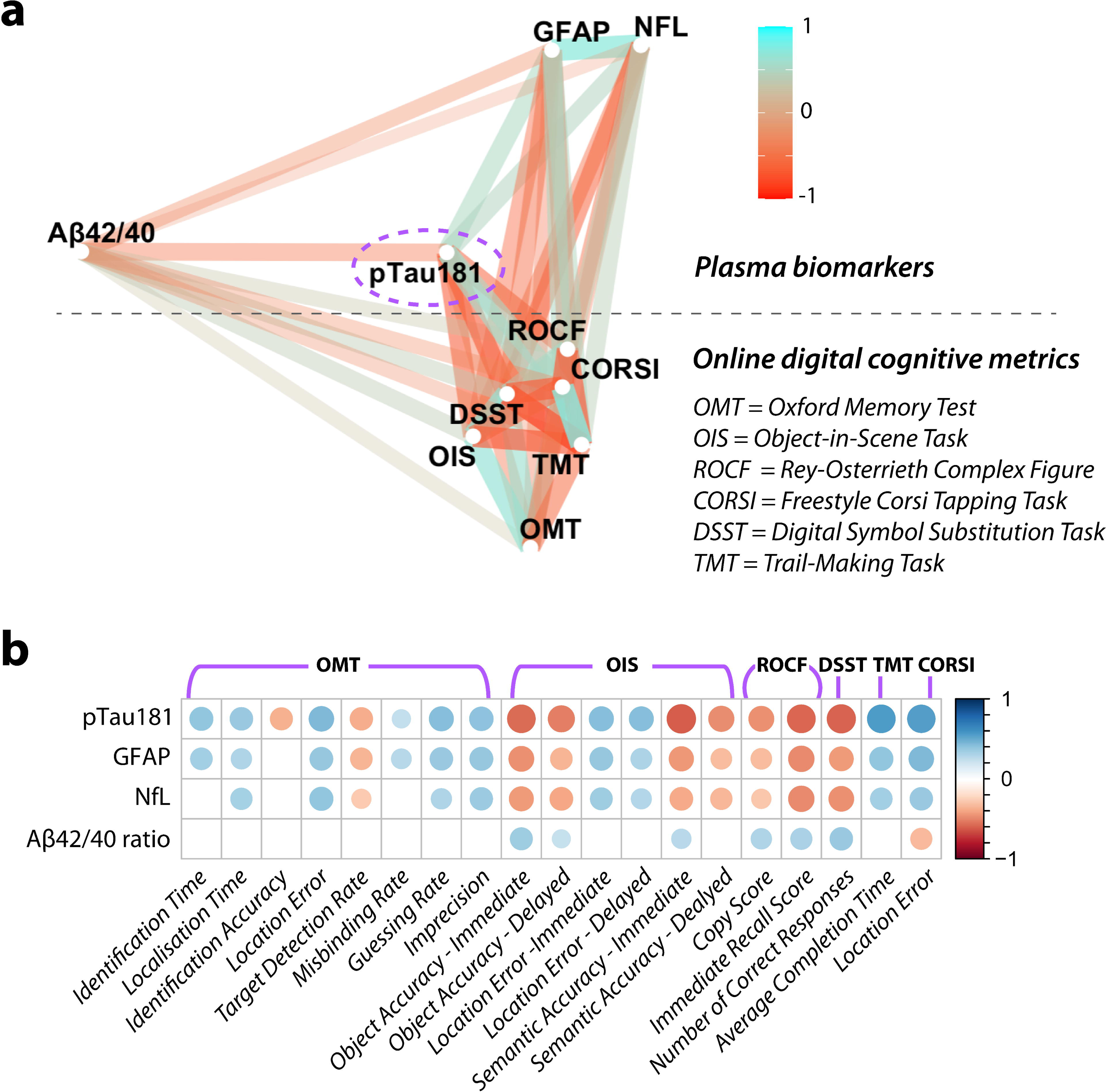
pTau181 shows the strongest relationship with digital cognitive tests (a) Network plot of relationships between all plasma biomarkers (clustered on the left) and online digital cognitive tasks (clustered on the right). Associations are presented in graded colours, where red is associated with a negative correlation and light blue with a positive correlation. The shorter distance between two metrics indicates a stronger relationship (larger correlation coefficient). For online tasks, only one metric was selected per task, according to the highest effect size in discriminating between groups. **(b)** Strength of the correlation is given by diameter of the circle, with positive correlations in blue and negative in red. All displayed correlations are significant after Benjamini–Hochberg correction for multiple comparisons.

This pattern of relationships can also be appreciated when looking at the individual correlation between each pair of biomarkers and cognitive metrics (Figure 4b). Across the different tasks examined, multiple metrics of short-term memory were correlated with pTau181, GFAP and NfL levels; the better the performance, the lower the levels of these three plasma biomarkers. Similarly, these plasma levels were also correlated with executive function metrics such as DSST and TMT, and with visuospatial ability as indicated by the ROCF copy score. In contrast, Aβ42/40 ratio was only weakly associated with selected short-term memory metrics and long-term memory metrics in OIS (e.g., immediate and delayed recall accuracy, immediate semantic accuracy) and performance at the Freestyle Corsi Block Task. Aβ42/40 ratio levels also correlated with visuospatial abilities (copy and immediate recall scores of ROCF) and processing speed (DSST). These results survived multiple comparison corrections across 76 correlations. Within group correlations between cognitive tests and plasma biomarkers did not survive corrections for multiple comparisons.

### 3.3 Which plasma/cognitive metric best predicts AD?

The selected variables were then ranked according to their importance in predicting group classification, i.e., AD or EHC (Figure 5a) using chi-square tests. The rank represents the negative log of the p-values. In this sample, all cognitive metrics ranked higher than plasma biomarkers. All tests and biomarkers were significant predictors of the group (all p < 0.001 except Aβ42/40 ratio, p = 0.002).

**Figure 5.**
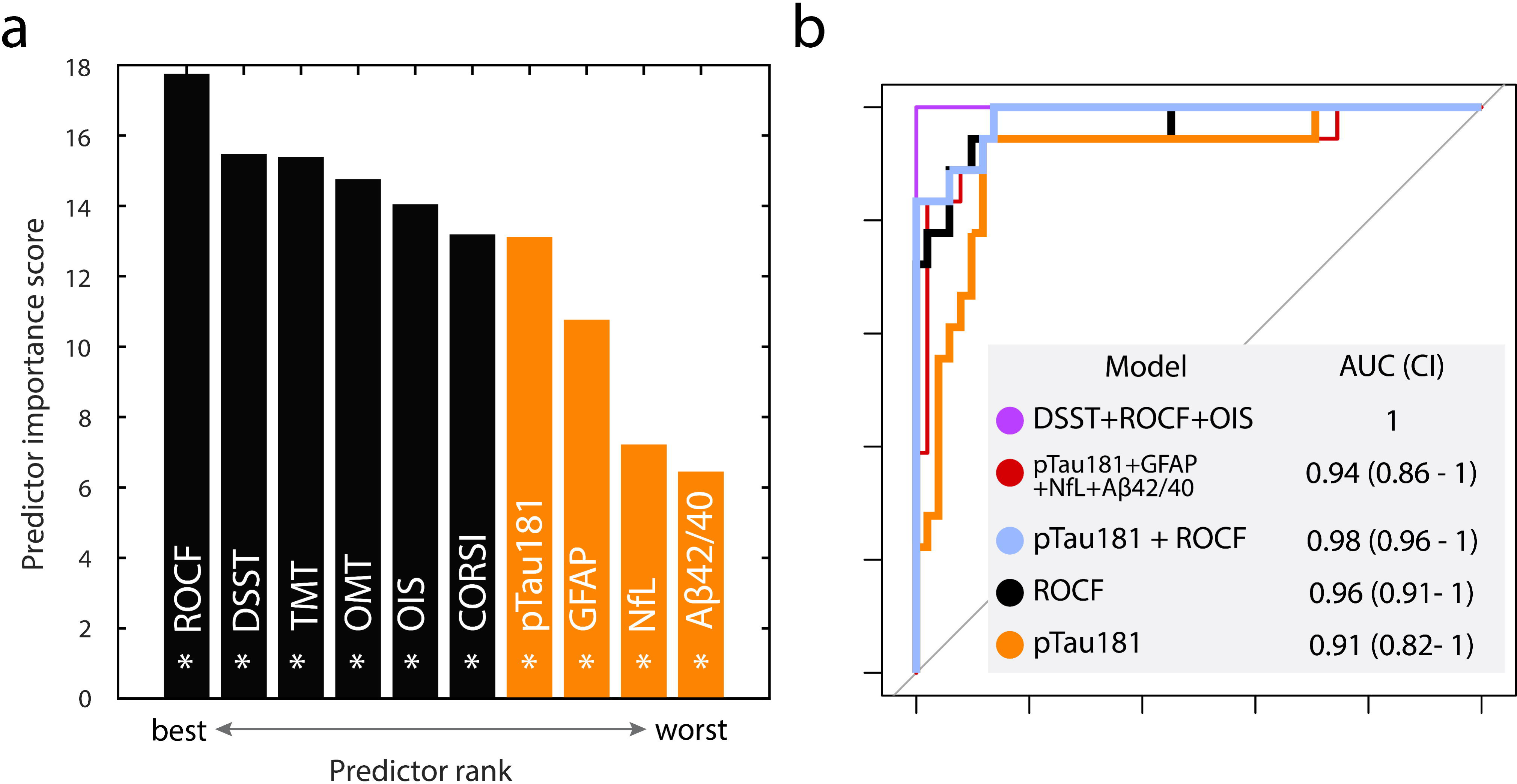
Which cognitive metric or plasma biomarker best predicts AD? (a) Ranked biomarkers and digital cognitive metrics in predicting group AD or control. Plasma biomarkers are marked in orange, while online cognitive metrics are marked in black. All metrics were significant in group classification. OMT = Identification accuracy of the Oxford Memory Task, OIS = Object Identification Accuracy in Immediate Recall of the Object-in-Scene Memory Task, ROCF = recall of the Rey–Osterrieth Complex Figure, DSST = number of correct responses of the Digit Symbol Substitution Task, TMT = average reaction time of the Trail Making Test, CORSI = average location error of the Freestyle Corsi Block Task. **(b)** ROC curves for group classification. Light blue shows the combined model with ROCF and pTau, with black line indicates ROCF alone model and orange shows the model pTau alone. The winning model, with ROCF, DSST and OIS, is displayed in a thinner, purple line. The model with the combination of all plasma biomarkers is depicted in a thinner, red line. AUC = area under the curve, CI = confidence interval.

We then explored which were the best predictors of tau and amyloid pathology, indexed respectively by pTau181 and the Aβ42/40 ratio. Most of our digital cognitive tests, except OMT, were better predictors of pTau181 levels compared to other plasma biomarkers such as GFAP and the Aβ42/40 ratio (Supplementary Figure 4a). The best-performing test in predicting pTau181 levels was DSST, followed by OIS, TMT and ROCF (all p < 0.001). All plasma biomarkers were significant predictors of pTau181 levels; NFL (p < 0.001), GFAP (p < 0.001), Aβ42/40 (p = 0.002).

Conversely, pTau181 was the only statistically significant predictor of amyloid burden (p = 0.018), (Supplementary Figure 4b). The best performing digital cognitive test to predict amyloid burden was DSST, but it was not statistically significant (p = 0.092).

Model comparison using the MuMIn R function was then used to choose the best combination of plasma biomarkers and cognitive metrics in predicting group classification (using logistic regression), pTau181 and the Aβ42/40 ratio levels (linear regression). The best model for predicting groups (the one with the lowest (AIC)), consisted of recall of the ROCF, Immediate Object Accuracy of the OIS and DSST (Figure 5b and Supplementary Figure 4c), with an AUC of 1. When Lasso penalization was introduced to avoid perfect separation of the best model, it still performed significantly well (AUC = 0.93).

If taken individually, the recall of the ROCF had an AUC of 0.962, DSST had an AUC of 0.955, and Immediate Object Accuracy of the OIS had an AUC of 0.937, while pTau181 had an AUC of 0.911 in predicting group classification. While ROCF alone was not statistically significantly different from the best model (ROCF: Z = 1.43, p = 0.15), the model which incorporated DSST, ROCF and OIS was better than a model containing only pTau181: Z = 2.0, p = 0.04). However, the model containing only ROCF was not better compared to the model with pTau181 alone (Z = 1.00, p = 0.31).

Of course, it is not surprising in this case that three cognitive tests together (DSST, ROCF and OIS) perform so well in distinguishing AD from healthy controls because of the separation in cognitive performance between these groups, as would be expected to be the case with any comparison between a group with established neurodegeneration and healthy people. What is of greater interest is how well combinations of plasma biomarkers of AD and cognitive test performance fare in making this discrimination. If pTau181 and ROCF are combined, this combination achieved an AUC of 0.983, and there was a non-significant trend towards the combination being statistically superior to pTau181 alone (Z = -1.86, p = 0.06). If all digital cognitive tests are combined the model achieved an AUC of 1, whilst if all plasma biomarkers are combined that resulted in an AUC of 0.940. These were however not statistically different (Z = -1.41, p-value = 0.16).

In comparison, ACE had an AUC of 0.97 in discriminating between groups, which was, however, not different compared to the best performing digital metric, ROCF (Z = 0.99, p-value = 0.82), nor to the best model (Z = -0.23, p-value = 0.32).

The best model for predicting pTau181 levels consisted of Aβ42/40, OIS, ROCF, DSST and OMT, which had an adjusted R^2^ of 0.50 (Supplementary Figure 4c). If single metrics were evaluated, in predicting pTau181 levels, ROCF, OIS and DSST had an adjusted R^2^ of respectively 0.31, 0.34 and 0.38, while in comparison, ACE had an adjusted R^2^ of 0.24. AICs were -7.55 (ROCF), -10.78 (OIS), -15.22 (DSST), and -1.2 (ACE), with the best-performing model being the one containing DSST (more negative). The winning model for predicting Aβ42/40 levels consisted only of pTau181, but had an overall poor model fit with an adjusted R^2^ of 0.14 (Supplementary Figure 4c).

## 4 Discussion

Currently, there are no published studies, to the best of our knowledge, which have reported on the relationship between performance on a panel of digital cognitive tests and plasma biomarkers of AD. The findings reported here demonstrate that patients with AD have impaired performance on our digital cognitive tests and that this is closely related to pathological blood-based biomarkers of the disease (Figures 3 and 4). Levels of plasma pTau181, GFAP and NfL were all highly correlated with several cognitive metrics, with pTau181 being the biomarker that showed the closest association to digital cognitive performance (Figure 4a). Overall, the results of this study show that it is feasible to deploy digital cognitive testing in AD, which would have the potential to make a significant impact on several fronts. Screening for the disease, recruitment and stratification into clinical trials, and longitudinal follow-up in intervention studies could all be transformed if cognitive testing were to be conducted robustly, remotely, and frequently [5]. Digital cognitive testing could make this happen.

Digital platforms are emerging as potential screening and diagnostic tools for people at risk of developing AD [50]. Most studies using such platforms have focused on screening healthy individuals [51]. Moreover, biomarker validation on these digital platforms is mostly limited to one single marker, frequently amyloid PET [51]. Some brief digital screening tools have demonstrated promise in differentiating amyloid-positive and tau-positive MCI patients (as measured by amyloid and tau PET) from MCI without evidence of amyloid or tau accumulation, but they have not been very good at separating healthy controls from people with MCI or prodromal AD [52]. A shorter digital version of the Face Name Associative Memory Exam (FNAME) or FACEmemory®, measuring episodic memory, has also been found to correlate with CSF levels of pTau181 and the Aβ42/40 [53]. Reduced learning over time over multiple exposures of the pen-and-paper FNAME has also been associated with amyloid and tau burden at PET [54] and the Spanish version of the same test (S-FNAME) has been shown not to be significantly correlated with plasma Aβ42/40 levels in a cohort of cognitively unimpaired individuals and patients with subjective cognitive impairment, while its composite score (SNF-F) showed a mild correlation (rho = 0.193, p = 0.006) in the same study [55]. However, the performance of the online shorter version of these tests compared to plasma biomarkers of AD is currently unknown.

One of the strengths of the current study is the inclusion of tests measuring different cognitive domains and the use of different plasma biomarkers, measuring not only amyloid and tau accumulation but also neuroinflammation and neurodegeneration. To our knowledge, no published study has previously investigated the relationship between these four biomarkers and performance on a digital platform in a mixed population of elderly healthy controls and patients with AD.

A key finding of this study is that digital cognitive metrics were more tightly correlated with pTau181 than the Aβ42/40 ratio (Figure 4 and Supplementary Figure 4). This is not entirely surprising, as amyloid burden has been shown to have a weaker association with cognition compared to tau [56,57]. The biological underpinning of such dissociation between small-magnitude correlations between amyloid plaques’ deposition and cognition in contrast to strong association with tau burden might be explained by the more tight association between tau accumulation, neuronal loss and subsequent gray matter atrophy [58], as opposed to a relatively slow, indolent accumulation of amyloid burden over time [59].

One possible explanation for the better performance of pTau181 compared to the Aβ42/40 ratio lies in the fact that pTau181 in blood correlates well with both amyloid and tau PET [10], and not only with amyloid burden. Moreover, unlike GFAP and NfL, pTau181 is AD-specific [11]. Whilst tau accumulation in the medial temporal lobe has often found to be associated with decline of episodic memory performance at standard pen-and-paper tests [58], it is also increasingly recognized that a decline in visual short-term memory performance at digital tasks, including one used in our battery, OMT, can be associated with medial temporal lobe atrophy in patients with AD [4]. Our results extend previous findings to biological hallmarks of AD measured by plasma biomarkers, and to other novel visual short-term memory tests, which perform better at a head-to-head comparison with the abovementioned test, OMT (Figure 5a).

A longitudinal increase in GFAP levels has been found to be associated with decline in memory, attention, and executive domains in cognitively unimpaired individuals at face-to-face tests, whilst in the same study an increase in NfL did now show such an association [60]. Our results show that these digital metrics, measuring visual short-term memory, long-term memory, visuospatial copying, executive function, and processing speed, are associated with both GFAP and NfL cross-sectional levels in a cohort of AD and HC, even if such associations were of lower magnitude compared to pTau181 (Figure 4a and 4b). Longitudinal studies are needed to see whether GFAP levels could be more tightly correlated compared to NfL to a decline of performance at these digital tasks in cognitively unimpaired controls.

Taking a closer look at single cognitive domains, a large meta-analysis that investigated different indices of amyloid positivity in cognitively unimpaired elderly adults without blood-based biomarkers showed that although episodic memory might be correlated with amyloid burden, global cognition and executive functions are not if assessed by amyloid PET [61]. On our digital platform, higher amyloid burden indexed by a lower Aβ42/40 ratio was not uniquely associated with long-term memory abilities but was correlated with performance across multiple tasks, measuring visual short-term and long-term memory, processing speed, and visuospatial function (Figure 4b). These results are encouraging, as they might suggest that these digital metrics are sensitive tools, showing a higher range of association with amyloid burden beyond episodic memory. However, similarly to pen-and-paper tests, these associations were of small magnitude and no associations were found in the sample of cognitively unimpaired individuals in this study after correcting for multiple comparisons across 76 correlations.

Notably, while performance on our digital cognitive tests was tightly associated with plasma biomarker levels, it was not statistically significantly correlated with measures of apathy (Supplementary Figure 3). Patients with AD are often reported to have higher rates of depression and apathy compared to age-matched healthy controls, a finding that was also present in our sample (Table 1). This supports the proposal that these metrics are related to cognitive performance and do not merely measure willingness to engage in the online task, which is a potential confounder. In this sample, depression was weakly associated with performance at our cognitive tests, but clustered further away on the right from most plasma and cognitive biomarkers (Supplementary Figure 3).

Crucially, in this sample all digital metrics were better predictors of group classification (AD or EHC) compared to plasma biomarkers (Figure 5a). In determining group classification using logistic regression, three digital tests performed best, as shown by the best model (lowest AIC) automatically selected (via the MumIn package) using model comparison of all possible combinations of digital and plasma biomarkers (see Supplementary Figure 4c). These were the recall on the ROCF test (a measure of visual episodic memory), Immediate Object Accuracy on the OIS task (a measure of visual short-term memory) and DSST (a measure of visual short-term memory). If these three tests were used in combination, this best-performing model had AUC of 1, achieving perfect separation of patients from controls (Figure 5b), which was better than the model containing pTau181. However, the best-performing digital metric, recall of ROCF, if used in isolation performed equally well as pTau181 in group classification. A biological confirmation is needed to correctly identify patients with AD, and our cognitive metrics should not be considered as a substitute for biological characterization. However, it is encouraging to observe that the combination of all our digital metrics performed equally as good as the combination of four different plasma biomarkers, and that the winning model for group classification contained three digital metrics and no plasma biomarker. Regarding prediction of pTau181 levels, DSST was the digital test which achieved the highest performance. We consider this result to be valuable as this test only takes 2 minutes to be performed, and might suggest that lengthy cognitive tests might not be necessarily needed to capture subtle biological variations.

Importantly, amongst the digital metrics, ROCF had comparable performance to the standard cognitive scores used (ACE) in group classification, and ROCF, OIS and DSST had better performance (higher adjusted R^2^ and lower AIC) than ACE in predicting pTau181 levels. This is encouraging, as in the future, the combination of these measures might be used as a proxy for standard cognitive metrics while saving a considerable amount of time in face-to-face appointments.

### Limitations and future directions

There are also several limitations to this study. One is that most of EHC1 and patients with AD came from a white Caucasian background, are highly educated and based in South-East England. We tried to address this by enrolling a second independent dataset of EHC2 to gather additional information regarding online performance in a more representative sample. However, our findings should be replicated in other populations with greater ethnic and socioeconomic diversity and wider geographical spread.

The very high AUCs of these metrics in predicting groups in this small, highly selected sample might partially explain the lack of positive contribution of adding cognition to plasma biomarkers. With accuracy being at ceiling, further evidence is needed to establish whether combining pTau181 to digital metrics might be beneficial in a larger dataset including different populations such as individuals with SCI or MCI. A bigger sample would also be required to assess the performance of these metrics in people with preclinical AD versus amyloid and tau-negative cognitively unimpaired healthy controls. Therefore, whether the combination of digital cognitive metrics and plasma biomarkers can be useful to stratify which individuals in the preclinical or prodromal phase of AD might be at risk of developing AD dementia remains to be established. However, it is encouraging that, even with a relatively small sample size, these metrics show a good correlation with several plasma biomarkers, surviving multiple comparisons and corrections for age, gender, and education, which are major confounders in both plasma biomarkers and cognitive assessments [62,63].

The patient population included in this study was already largely at the AD dementia stage, where cognitive impairment is overt. In this sample, plasma pTau181 was the biomarker that was more closely associated with cognitive metrics and the best predictor of group classification. However, we cannot exclude that other biomarkers such as pTau217 could show an even higher association with cross-sectional or longitudinal cognitive function in the same population, as some evidence suggests [64,65]. Also, early markers of amyloid deposition such as pTau217, pTau213 and GFAP may be more closely linked with cognitive changes in the early phases of the disease [14,15,64,65].

Finally, a limitation of remote testing is the lower level of control over experimental conditions at home compared to testing in hospital settings. In this study, patients with AD had different levels of cognitive impairment, and some patients needed external support to ensure they understood task instructions and could carry out the task. This support was delivered by the caregiver if tests were performed at home, or by a member of the team if the tests were done at the hospital. However, these are intrinsic limitations of remote testing and are not unique to our study.

### Conclusion

To conclude, digital cognitive metrics were tightly associated with several plasma biomarkers, particularly pTau181, but also with GFAP and NfL, and to a much lesser extent with the Aβ42/40 ratio. Adding these metrics to pTau181 did not improve group classification in this sample, but the best performing metric, the recall of ROCF, performed at par with pTau181 levels. As plasma biomarkers are being proposed as equivalent to CSF biomarkers in the forthcoming NIA-AA revised criteria for AD [66], and given their increased use in clinical practice [67], implementation of a digital cognitive platform that has been validated with AD plasma biomarkers provides an important step forward for future large-scale deployment.

## Data availability

De-identified data supporting this study may be shared based on reasonable written requests to the corresponding author. Access to de-identified data will require a Data Access Agreement and IRB clearance, which will be considered by the institutions who provided the data for this research.

## Code Availability

The source code will be shared using a Creative Commons NC-ND 4.0 international licence upon reasonable written request to the corresponding author and requires a research use agreement.

## Authors contribution

STo: conceptualization, participants’ recruitment, project administration, resources management, data collection and curation, analysis, data visualisation, writing, SZ: conceptualization, software development, code development, digital platform data curation and management, analysis, data visualisation, writing, AS: cognitive testing data collection and curation, BA: biomarker samples preprocessing and data curation, AG: cognitive testing data collection and curation, AH: biomarker samples analysis, STh and SM: participants’ recruitment, HZ: biomarker samples analysis, MH: conceptualization, supervision, resources management, funding acquisition, participants’ recruitment, writing. All authors read and approved the final manuscript.

## Acknowledgements and funding

This research was supported by funding from the Wellcome Trust and NIHR Oxford Health Biomedical Research Centre. S.T., S.Z. and M.H. were funded by the Wellcome Trust (206330/Z/17/Z). The project was further supported by a Guarantors of Brain post-doctoral fellowship to S.T.. H.Z. is a Wallenberg Scholar supported by grants from the Swedish Research Council (#2022-01018 and #2019-02397), the European Union’s Horizon Europe research and innovation programme under grant agreement No 101053962, Swedish State Support for Clinical Research (#ALFGBG-71320), the Alzheimer Drug Discovery Foundation (ADDF), USA (#201809-2016862), the AD Strategic Fund and the Alzheimer’s Association (#ADSF-21-831376-C, #ADSF-21-831381-C, and #ADSF-21-831377-C), the Bluefield Project, the Olav Thon Foundation, the Erling-Persson Family Foundation, Stiftelsen för Gamla Tjänarinnor, Hjärnfonden, Sweden (#FO2022-0270), the European Union’s Horizon 2020 research and innovation programme under the Marie Skłodowska-Curie grant agreement No 860197 (MIRIADE), the European Union Joint Programme – Neurodegenerative Disease Research (JPND2021-00694), the National Institute for Health and Care Research University College London Hospitals Biomedical Research Centre, and the UK Dementia Research Institute at UCL (UKDRI-1003). The funder played no role in study design, data collection, analysis and interpretation of data, or the writing of this manuscript.

## References

[1] Sims JR, Zimmer JA, Evans CD, Lu M, Ardayfio P, Sparks J, et al. Donanemab in Early Symptomatic Alzheimer Disease: The TRAILBLAZER-ALZ 2 Randomized Clinical Trial. JAMA 2023. 10.1001/jama.2023.13239.

[2] van Dyck CH, Swanson CJ, Aisen P, Bateman RJ, Chen C, Gee M, et al. Lecanemab in Early Alzheimer’s Disease. New England Journal of Medicine 2023;388:9–21. 10.1056/NEJMoa2212948.

[3] Jack CR, Knopman DS, Jagust WJ, Shaw LM, Aisen PS, Weiner MW, et al. Hypothetical model of dynamic biomarkers of the Alzheimer’s pathological cascade. Lancet Neurol 2010;9:119–28. 10.1016/S1474-4422(09)70299-6.

[4] Liang Y, Pertzov Y, Nicholas JM, Henley SMD, Crutch S, Woodward F, et al. Visual short-term memory binding deficit in familial Alzheimer’s disease. Cortex 2016;78:150–64. 10.1016/j.cortex.2016.01.015.

[5] Jutten RJ, Papp KV, Hendrix S, Ellison N, Langbaum JB, Donohue MC, et al. Why a clinical trial is as good as its outcome measure: A framework for the selection and use of cognitive outcome measures for clinical trials of Alzheimer’s disease. Alzheimers Dement 2023;19:708–20. 10.1002/alz.12773.

[6] Doecke JD, Pérez-Grijalba V, Fandos N, Fowler C, Villemagne VL, Masters CL, et al. Total Aβ42/Aβ40 ratio in plasma predicts amyloid-PET status, independent of clinical AD diagnosis. Neurology 2020;94:e1580–91. 10.1212/WNL.0000000000009240.

[7] Giudici KV, de Souto Barreto P, Guyonnet S, Li Y, Bateman RJ, Vellas B, et al. Assessment of Plasma Amyloid-β42/40 and Cognitive Decline Among Community-Dwelling Older Adults. JAMA Network Open 2020;3:e2028634. 10.1001/jamanetworkopen.2020.28634.

[8] Stockmann J, Verberk IMW, Timmesfeld N, Denz R, Budde B, Lange-Leifhelm J, et al. Amyloid-β misfolding as a plasma biomarker indicates risk for future clinical Alzheimer’s disease in individuals with subjective cognitive decline. Alzheimer’s Research & Therapy 2020;12:169. 10.1186/s13195-020-00738-8.

[9] Verberk IMW, Hendriksen HMA, van Harten AC, Wesselman LMP, Verfaillie SCJ, van den Bosch KA, et al. Plasma amyloid is associated with the rate of cognitive decline in cognitively normal elderly: the SCIENCe project. Neurobiol Aging 2020;89:99–107. 10.1016/j.neurobiolaging.2020.01.007.

[10] Janelidze S, Mattsson N, Palmqvist S, Smith R, Beach TG, Serrano GE, et al. Plasma P-tau181 in Alzheimer’s disease: relationship to other biomarkers, differential diagnosis, neuropathology and longitudinal progression to Alzheimer’s dementia. Nat Med 2020;26:379–86. 10.1038/s41591-020-0755-1.

[11] Karikari TK, Pascoal TA, Ashton NJ, Janelidze S, Benedet AL, Rodriguez JL, et al. Blood phosphorylated tau 181 as a biomarker for Alzheimer’s disease: a diagnostic performance and prediction modelling study using data from four prospective cohorts. Lancet Neurol 2020;19:422–33. 10.1016/S1474-4422(20)30071-5.

[12] Mielke MM, Hagen CE, Xu J, Chai X, Vemuri P, Lowe VJ, et al. Plasma phospho-tau181 increases with Alzheimer’s disease clinical severity and is associated with tau- and amyloid-positron emission tomography. Alzheimers Dement 2018;14:989–97. 10.1016/j.jalz.2018.02.013.

[13] Moscoso A, Grothe MJ, Ashton NJ, Karikari TK, Rodriguez JL, Snellman A, et al. Time course of phosphorylated-tau181 in blood across the Alzheimer’s disease spectrum. Brain 2021;144:325–39. 10.1093/brain/awaa399.

[14] Chatterjee P, Pedrini S, Stoops E, Goozee K, Villemagne VL, Asih PR, et al. Plasma glial fibrillary acidic protein is elevated in cognitively normal older adults at risk of Alzheimer’s disease. Transl Psychiatry 2021;11:1–10. 10.1038/s41398-020-01137-1.

[15] Cicognola C, Janelidze S, Hertze J, Zetterberg H, Blennow K, Mattsson-Carlgren N, et al. Plasma glial fibrillary acidic protein detects Alzheimer pathology and predicts future conversion to Alzheimer dementia in patients with mild cognitive impairment. Alzheimer’s Research & Therapy 2021;13:68. 10.1186/s13195-02100804-9.

[16] Pereira JB, Janelidze S, Smith R, Mattsson-Carlgren N, Palmqvist S, Teunissen CE, et al. Plasma GFAP is an early marker of amyloid-β but not tau pathology in Alzheimer’s disease. Brain 2021;144:3505–16. 10.1093/brain/awab223.

[17] Verberk IMW, Thijssen E, Koelewijn J, Mauroo K, Vanbrabant J, de Wilde A, et al. Combination of plasma amyloid beta(1-42/1-40) and glial fibrillary acidic protein strongly associates with cerebral amyloid pathology. Alzheimer’s Research & Therapy 2020;12:118. 10.1186/s13195-020-00682-7.

[18] Abdelhak A, Foschi M, Abu-Rumeileh S, Yue JK, D’Anna L, Huss A, et al. Blood GFAP as an emerging biomarker in brain and spinal cord disorders. Nat Rev Neurol 2022;18:158–72. 10.1038/s41582-021-00616-3.

[19] Benedet AL, Ashton NJ, Pascoal TA, Leuzy A, Mathotaarachchi S, Kang MS, et al. Plasma neurofilament light associates with Alzheimer’s disease metabolic decline in amyloid-positive individuals. Alzheimers Dement (Amst) 2019;11:679–89. 10.1016/j.dadm.2019.08.002.

[20] Marks JD, Syrjanen JA, Graff-Radford J, Petersen RC, Machulda MM, Campbell MR, et al. Comparison of plasma neurofilament light and total tau as neurodegeneration markers: associations with cognitive and neuroimaging outcomes. Alzheimer’s Research & Therapy 2021;13:199. 10.1186/s13195-021-00944-y.

[21] Mattsson N, Cullen NC, Andreasson U, Zetterberg H, Blennow K. Association Between Longitudinal Plasma Neurofilament Light and Neurodegeneration in Patients With Alzheimer Disease. JAMA Neurology 2019;76:791–9. 10.1001/jamaneurol.2019.0765.

[22] Palmqvist S, Tideman P, Cullen N, Zetterberg H, Blennow K, Dage JL, et al. Prediction of future Alzheimer’s disease dementia using plasma phospho-tau combined with other accessible measures. Nat Med 2021;27:1034–42. 10.1038/s41591-021-01348-z.

[23] Karikari TK, Ashton NJ, Brinkmalm G, Brum WS, Benedet AL, Montoliu-Gaya L, et al. Blood phospho-tau in Alzheimer disease: analysis, interpretation, and clinical utility. Nat Rev Neurol 2022;18:400–18. 10.1038/s41582-022-00665-2.

[24] Mielke MM, Frank RD, Dage JL, Jeromin A, Ashton NJ, Blennow K, et al. Comparison of Plasma Phosphorylated Tau Species With Amyloid and Tau Positron Emission Tomography, Neurodegeneration, Vascular Pathology, and Cognitive Outcomes. JAMA Neurol 2021;78:1108–17. 10.1001/jamaneurol.2021.2293.

[25] O’Connor A, Karikari TK, Poole T, Ashton NJ, Lantero Rodriguez J, Khatun A, et al. Plasma phospho-tau181 in presymptomatic and symptomatic familial Alzheimer’s disease: a longitudinal cohort study. Mol Psychiatry 2021;26:5967–76. 10.1038/s41380-020-0838-x.

[26] Mattsson-Carlgren N, Janelidze S, Palmqvist S, Cullen N, Svenningsson AL, Strandberg O, et al. Longitudinal plasma p-tau217 is increased in early stages of Alzheimer’s disease. Brain 2020;143:3234–41. 10.1093/brain/awaa286.

[27] Donohue MC, Sperling RA, Salmon DP, Rentz DM, Raman R, Thomas RG, et al. The preclinical Alzheimer cognitive composite: measuring amyloid-related decline. JAMA Neurol 2014;71:961–70. 10.1001/jamaneurol.2014.803.

[28] Oeckl P, Halbgebauer S, Anderl-Straub S, Steinacker P, Huss AM, Neugebauer H, et al. Glial Fibrillary Acidic Protein in Serum is Increased in Alzheimer’s Disease and Correlates with Cognitive Impairment. J Alzheimers Dis 2019;67:481–8. 10.3233/JAD-180325.

[29] Ashton NJ, Janelidze S, Al Khleifat A, Leuzy A, van der Ende EL, Karikari TK, et al. A multicentre validation study of the diagnostic value of plasma neurofilament light. Nat Commun 2021;12:3400. 10.1038/s41467-021-23620-z.

[30] Rajan KB, Aggarwal NT, McAninch EA, Weuve J, Barnes LL, Wilson RS, et al. Remote Blood Biomarkers of Longitudinal Cognitive Outcomes in a Population Study. Ann Neurol 2020;88:1065–76. 10.1002/ana.25874.

[31] Mielke MM, Syrjanen JA, Blennow K, Zetterberg H, Vemuri P, Skoog I, et al. Plasma and CSF neurofilament light: Relation to longitudinal neuroimaging and cognitive measures. Neurology 2019;93:e252–60. 10.1212/WNL.0000000000007767.

[32] Moscoso A, Grothe MJ, Ashton NJ, Karikari TK, Lantero Rodríguez J, Snellman A, et al. Longitudinal Associations of Blood Phosphorylated Tau181 and Neurofilament Light Chain With Neurodegeneration in Alzheimer Disease. JAMA Neurology 2021;78:396–406. 10.1001/jamaneurol.2020.4986.

[33] Mattsson-Carlgren N, Salvadó G, Ashton NJ, Tideman P, Stomrud E, Zetterberg H, et al. Prediction of Longitudinal Cognitive Decline in Preclinical Alzheimer Disease Using Plasma Biomarkers. JAMA Neurology 2023;80:360–9. 10.1001/jamaneurol.2022.5272.

[34] McKhann GM, Knopman DS, Chertkow H, Hyman BT, Jack CR, Kawas CH, et al. The diagnosis of dementia due to Alzheimer’s disease: recommendations from the National Institute on Aging-Alzheimer’s Association workgroups on diagnostic guidelines for Alzheimer’s disease. Alzheimers Dement 2011;7:263–9. 10.1016/j.jalz.2011.03.005.

[35] Mathuranath PS, Nestor PJ, Berrios GE, Rakowicz W, Hodges JR. A brief cognitive test battery to differentiate Alzheimer’s disease and frontotemporal dementia. Neurology 2000;55:1613–20. 10.1212/01.wnl.0000434309.85312.19.

[36] Henrich J, Heine SJ, Norenzayan A. The weirdest people in the world? Behav Brain Sci 2010;33:61–83; discussion 83-135. 10.1017/S0140525X0999152X.

[37] Fandos N, Pérez-Grijalba V, Pesini P, Olmos S, Bossa M, Villemagne VL, et al. Plasma amyloid β 42/40 ratios as biomarkers for amyloid β cerebral deposition in cognitively normal individuals. Alzheimers Dement (Amst) 2017;8:179–87. 10.1016/j.dadm.2017.07.004.

[38] Ashton NJ, Leuzy A, Lim YM, Troakes C, Hortobágyi T, Höglund K, et al. Increased plasma neurofilament light chain concentration correlates with severity of post-mortem neurofibrillary tangle pathology and neurodegeneration. Acta Neuropathologica Communications 2019;7:5. 10.1186/s40478-018-0649-3.

[39] Pavisic IM, Nicholas JM, Pertzov Y, O’Connor A, Liang Y, Collins JD, et al. Visual short-term memory impairments in presymptomatic familial Alzheimer’s disease: A longitudinal observational study. Neuropsychologia 2021;162:108028. 10.1016/j.neuropsychologia.2021.108028.

[40] Pertzov Y, Miller TD, Gorgoraptis N, Caine D, Schott JM, Butler C, et al. Binding deficits in memory following medial temporal lobe damage in patients with voltage-gated potassium channel complex antibody-associated limbic encephalitis. Brain 2013;136:2474–85. 10.1093/brain/awt129.

[41] Li Q, Joo SJ, Yeatman JD, Reinecke K. Controlling for Participants’ Viewing Distance in Large-Scale, Psychophysical Online Experiments Using a Virtual Chinrest. Sci Rep 2020;10:904. 10.1038/s41598-019-57204-1.

[42] Osterrieth PA. Le test de copie d’une figure complexe; contribution à l’étude de la perception et de la mémoire. [Test of copying a complex figure; contribution to the study of perception and memory.]. Archives de Psychologie 1944;30:206–356.

[43] Bowie CR, Harvey PD. Administration and interpretation of the Trail Making Test. Nat Protoc 2006;1:2277–81. 10.1038/nprot.2006.390.

[44] Sherman E, Tan J, Hrabok and M. A Compendium of Neuropsychological Tests: Fundamentals of Neuropsychological Assessment and Test Reviews for Clinical Practice. Fourth Edition, Fourth Edition. Oxford, New York: Oxford University Press; 2023.

[45] Zeng Z, Miao C, Leung C, Shen Z. Computerizing Trail Making Test for long-term cognitive self-assessment. International Journal of Crowd Science 2017;1:83–99. 10.1108/IJCS-12-2016-0002.

[46] Corsi PM. Human memory and the medial temporal region of the brain. McGill University, 1972.

[47] Arce T, McMullen K. The Corsi Block-Tapping Test: Evaluating methodological practices with an eye towards modern digital frameworks. Computers in Human Behavior Reports 2021;4:100099. 10.1016/j.chbr.2021.100099.

[48] Ang Y-S, Lockwood P, Apps MAJ, Muhammed K, Husain M. Distinct Subtypes of Apathy Revealed by the Apathy Motivation Index. PLOS ONE 2017;12:e0169938. 10.1371/journal.pone.0169938.

[49] Yesavage J. Geriatric depression scale. Psychopharmacology Bulletin, The Clearinghouse; 1988, p. 709–11.

[50] Staffaroni AM, Tsoy E, Taylor J, Boxer AL, Possin KL. Digital Cognitive Assessments for Dementia. Pract Neurol (Fort Wash Pa) 2020;2020:24–45.

[51] Öhman F, Hassenstab J, Berron D, Schöll M, Papp KV. Current advances in digital cognitive assessment for preclinical Alzheimer’s disease. Alz & Dem Diag Ass & Dis Mo 2021;13. 10.1002/dad2.12217.

[52] Alden EC, Pudumjee SB, Lundt ES, Albertson SM, Machulda MM, Kremers WK, et al. Diagnostic accuracy of the Cogstate Brief Battery for prevalent MCI and prodromal AD (MCI A+T+) in a population-based sample. Alzheimer’s & Dementia 2021;17:584–94. 10.1002/alz.12219.

[53] Alegret M, Sotolongo-Grau O, de Antonio EE, Pérez-Cordón A, Orellana A, Espinosa A, et al. Automatized FACEmemory® scoring is related to Alzheimer’s disease phenotype and biomarkers in early-onset mild cognitive impairment: the BIOFACE cohort. Alzheimer’s Research & Therapy 2022;14:43. 10.1186/s13195-022-00988-8.

[54] Samaroo A, Amariglio RE, Burnham S, Sparks P, Properzi M, Schultz AP, et al. Diminished Learning Over Repeated Exposures (LORE) in preclinical Alzheimer’s disease. Alzheimers Dement (Amst) 2020;12:e12132. 10.1002/dad2.12132.

[55] Pascual-Lucas M, Allué JA, Sarasa L, Fandos N, Castillo S, Terencio J, et al. Clinical performance of an antibody-free assay for plasma Aβ42/Aβ40 to detect early alterations of Alzheimer’s disease in individuals with subjective cognitive decline. Alzheimer’s Research & Therapy 2023;15:2. 10.1186/s13195-022-01143-z.

[56] Giannakopoulos P, Herrmann FR, Bussière T, Bouras C, Kövari E, Perl DP, et al. Tangle and neuron numbers, but not amyloid load, predict cognitive status in Alzheimer’s disease. Neurology 2003;60:1495–500. 10.1212/01.wnl.0000063311.58879.01.

[57] Ossenkoppele R, Smith R, Mattsson-Carlgren N, Groot C, Leuzy A, Strandberg O, et al. Accuracy of Tau Positron Emission Tomography as a Prognostic Marker in Preclinical and Prodromal Alzheimer Disease: A Head-to-Head Comparison Against Amyloid Positron Emission Tomography and Magnetic Resonance Imaging. JAMA Neurology 2021;78:961–71. 10.1001/jamaneurol.2021.1858.

[58] Bejanin A, Schonhaut DR, La Joie R, Kramer JH, Baker SL, Sosa N, et al. Tau pathology and neurodegeneration contribute to cognitive impairment in Alzheimer’s disease. Brain 2017;140:3286–300. 10.1093/brain/awx243.

[59] Nelson PT, Alafuzoff I, Bigio EH, Bouras C, Braak H, Cairns NJ, et al. Correlation of Alzheimer Disease Neuropathologic Changes With Cognitive Status: A Review of the Literature. Journal of Neuropathology & Experimental Neurology 2012;71:362–81. 10.1097/NEN.0b013e31825018f7.

[60] Verberk IMW, Laarhuis MB, Bosch KA van den, Ebenau JL, Leeuwenstijn M van, Prins ND, et al. Serum markers glial fibrillary acidic protein and neurofilament light for prognosis and monitoring in cognitively normal older people: a prospective memory clinic-based cohort study. The Lancet Healthy Longevity 2021;2:e87–95. 10.1016/S2666-7568(20)30061-1.

[61] Hedden T, Oh H, Younger AP, Patel TA. Meta-analysis of amyloid-cognition relations in cognitively normal older adults. Neurology 2013;80:1341–8. 10.1212/WNL.0b013e31828ab35d.

[62] Berron D, Ziegler G, Vieweg P, Billette O, Güsten J, Grande X, et al. Feasibility of Digital Memory Assessments in an Unsupervised and Remote Study Setting. Front Digit Health 2022;4:892997. 10.3389/fdgth.2022.892997.

[63] Ferreira PCL, Zhang Y, Snitz B, Chang C-CH, Bellaver B, Jacobsen E, et al. Plasma biomarkers identify older adults at risk of Alzheimer’s disease and related dementias in a real-world population-based cohort. Alzheimer’s & Dementia 2023;n/a. 10.1002/alz.12986.

[64] Ashton NJ, Janelidze S, Mattsson-Carlgren N, Binette AP, Strandberg O, Brum WS, et al. Differential roles of Aβ42/40, p-tau231 and p-tau217 for Alzheimer’s trial selection and disease monitoring. Nat Med 2022;28:2555–62. 10.1038/s41591-022-02074-w.

[65] Milà-Alomà M, Ashton NJ, Shekari M, Salvadó G, Ortiz-Romero P, Montoliu-Gaya L, et al. Plasma p-tau231 and p-tau217 as state markers of amyloid-β pathology in preclinical Alzheimer’s disease. Nat Med 2022;28:1797–801. 10.1038/s41591-022-01925-w.

[66] NIA-AA Workshop. NIA-AA Revised Clinical Criteria for Alzheimer’s Disease, Amsterdam, Netherlands: 2023.

[67] Hampel H, Hu Y, Cummings J, Mattke S, Iwatsubo T, Nakamura A, et al. Blood-based biomarkers for Alzheimer’s disease: Current state and future use in a transformed global healthcare landscape. Neuron 2023:S0896627323003902. 10.1016/j.neuron.2023.05.017.

