## Supplementary data for "Relationship of plasma biomarkers to digital cognitive tests in alzheimer’s disease"

### Supplementary Materials

#### Supplementary Methods 1 | Socioeconomic status and ethnic group of EHC2 group

340 of the 352 EHC2 participants provided us with their ethnic group. The distribution of the EHC2 is closely aligned to the UK population, with a good coverage of four major ethnic groups [1].

**Table 1. Ethnic groups.**

"White" group includes Caucasian, Sephardic Jew, Mexican, Middle Eastern and Latino/Hispanic. "Asian" group include South Asian, East Asian and South East Asian. "Black" group include African Black and Caribbean Black. "Other" group include Mixed and other groups that were not listed above.

| Ethnicity | AD | EHC1 | EHC2 |
|---|---|---|---|
| White | 77.8% | 66.0% | 74.4% |
| Asian | 2.8% | 0.0% | 7.9% |
| Black | 0.0% | 0.0% | 10.3% |
| Other | 19.4% | 34.0% | 7.4% |
| Ethnicity unknown | 2.8% | 3.8% | 3.5% |

Subjective socioeconomic status, as measured by the MacArthur Scale of Subjective Social Status, is a typical technique to get an understanding of the participants' socioeconomic position without directly measuring the objective socioeconomic status (e.g., household income, property ownership, etc.). It depicts a ten-rung ladder, with participants asked to place themselves on one of the rungs by determining where they stand in relation to other people in their country [2] and has shown good test-retest reliability and is expected to be strongly associated with objective socioeconomic status [3]. The distribution of EHC2's subjective socioeconomic status is depicted in Figure 1 below.

**Figure 1. Distribution of subjective socioeconomic status in EHC2.**

204 of 352 participants provided their answer to the MacArthur Scale of Subjective Social Status. The group mean was 5.7 (1.5) which was within the expected range of the UK population.

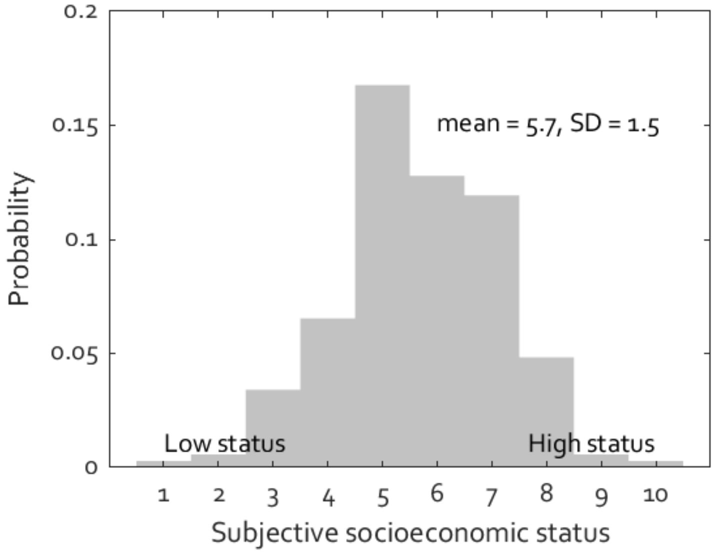

Even though an up-to-date distribution of subjective socioeconomic status in the UK is not publicly available, one way to estimate the average norm of subjective socioeconomic status is based on the countries with gross domestic product (GDP) per capita close to the UK. According to a recent social science report containing over 33,000 responses from 29 countries worldwide, GDP per capita is a strong predictor of average subjective socioeconomic status [4]. The GDP per capita of Sweden, the Netherlands, and Finland was very similar to that of the UK, and their average normative subjective socioeconomic status ranged between 5.6 and 6.0. The new EHC2 group's average socioeconomic level is 5.7 (SD=1.5), which falls within this expected range for the UK population. This, although indirect, implies that, in terms of socioeconomic position and ethnic groups, our EHC2 is a good general population representative.

#### Supplementary Table 1 | Digital cognitive metrics

The raw measures are followed by normalised measures.

| Digital Cognitive Tasks | Metrics | AD (n= 46) | EHC 1 (n= 53) | EHC 2 (n=352) | AD vs EHC 1 | AD vs EHC 2 | EHC 1 vs EHC 2 |
|---|---|---|---|---|---|---|---|
| OMT | Identification Accuracy (%) | 81.3 (10.1) | 92.3 (4.9) | 89.6 (7.7) | rrb = 0.63, p <0.0001 | rrb = 0.47, p = 0.0001 | rrb = -0.21, n.s. |
| | Identification Accuracy (z) | -1.3 (1.9) | 0.6 (1.0) | -0.2 (1.4) | rrb = 0.59, p <0.0001 | rrb = 0.38, n.s. | rrb = -0.37, p <0.0001 |
| | Location Error (pixels) | 227.5 (92.8) | 105.9 (43.6) | 85.9 (45.1) | rrb = -0.69, p <0.0001 | rrb = -0.81, p <0.0001 | rrb = -0.38, p <0.0001 |
| | Location Error (z) | 4.4 (3.4) | 0.9 (1.7) | 0.2 (1.4) | rrb = -0.64, p <0.0001 | rrb = -0.77, p <0.0001 | rrb = -0.33, p = 0.0002 |
| | Identification Time (s) | 4.1 (2.1) | 2.6 (1.3) | 2.4 (1.3) | rrb = -0.46, n.s. | rrb = -0.53, p <0.0001 | rrb = -0.24, n.s. |
| | Identification Time (z) | 3.0 (3.7) | 0.7 (2.4) | 0.3 (2.3) | rrb = -0.44, n.s. | rrb = -0.51, p <0.0001 | rrb = -0.19, n.s. |
| | Localisation Time (s) | 8.2 (3.5) | 5.4 (2.1) | 3.1 (1.5) | rrb = -0.49, p = 0.0007 | rrb = -0.89, p <0.0001 | rrb = -0.67, p <0.0001 |
| | Localisation Time (z) | 5.5 (4.6) | 2.5 (2.7) | 0.2 (1.4) | rrb = -0.43, n.s. | rrb = -0.87, p <0.0001 | rrb = -0.63, p <0.0001 |
| | Target Detection Rate (%) | 53.1 (17.0) | 76.2 (9.1) | 69.2 (13.8) | rrb = 0.74, p <0.0001 | rrb = 0.56, p <0.0001 | rrb = -0.34, p = 0.0001 |
| | Target Detection Rate (z) | -1.3 (1.6) | 0.7 (0.9) | -0.1 (1.2) | rrb = 0.71, p <0.0001 | rrb = 0.47, p = 0.0002 | rrb = -0.46, p <0.0001 |
| | Misbinding Error Rate (%) | 36.9 (11.8) | 21.7 (7.9) | 28.1 (10.2) | rrb = -0.68, p <0.0001 | rrb = -0.47, p = 0.0001 | rrb = 0.42, p <0.0001 |
| | Misbinding Error Rate(z) | 0.7 (1.4) | -1.0 (0.9) | -0.1 (1.1) | rrb = -0.67, p <0.0001 | rrb = -0.40, n.s. | rrb = 0.50, p <0.0001 |
| | Guessing Error Rate (%) | 28.4 (12.2) | 13.0 (5.9) | 16.7 (9.6) | rrb = -0.73, p <0.0001 | rrb = -0.57, p <0.0001 | rrb = 0.25, n.s. |
| | Guessing Error Rate (z) | 1.6 (1.8) | -0.5 (0.9) | 0.2 (1.3) | rrb = -0.71, p <0.0001 | rrb = -0.49, p = 0.0001 | rrb = 0.35, p <0.0001 |

| | | | | | | | |
|---|---|---|---|---|---|---|---|
| | Imprecision (pixels) | 133.3 (67.4) | 67.4 (27.9) | 54.2 (27.0) | rrb = -0.63, p <0.0001 | rrb = -0.76, p <0.0001 | rrb = -0.39, p <0.0001 |
| | Imprecision (z) | 5.1 (4.2) | 1.4 (2.4) | 0.2 (1.4) | rrb = -0.57, p = 0.0001 | rrb = -0.76, p <0.0001 | rrb = -0.44, p <0.0001 |
| **OIS** | Object Identification Accuracy - Immediate Recall (%) | 74.3 (16.1) | 95.5 (5.3) | 95.0 (7.5) | rrb = 0.87, p <0.0001 | rrb = 0.83, p <0.0001 | rrb = 0.05, n.s. |
| | Object Identification Accuracy - Immediate Recall (z) | -3.8 (3.2) | 0.0 (1.2) | -0.3 (1.5) | rrb = 0.85, p <0.0001 | rrb = 0.77, p <0.0001 | rrb = -0.11, n.s. |
| | Object Identification Accuracy - Delayed Recall (%) | 28.9 (19.9) | 68.4 (18.2) | 68.9 (18.7) | rrb = 0.83, p <0.0001 | rrb = 0.84, p <0.0001 | rrb = 0.01, n.s. |
| | Object Identification Accuracy - Delayed Recall (z) | -2.5 (1.3) | 0.0 (1.2) | -0.1 (1.1) | rrb = 0.82, p <0.0001 | rrb = 0.82, p <0.0001 | rrb = -0.07, n.s. |
| | Semantic Identification Accuracy - Immediate Recall (%) | 90.9 (10.8) | 99.6 (1.5) | 99.5 (3.4) | rrb = 0.64, p <0.0001 | rrb = 0.65, p <0.0001 | rrb = 0.06, n.s. |
| | Semantic Identification Accuracy - Immediate Recall (z) | -5.7 (4.6) | -0.6 (2.8) | -0.7 (4.0) | rrb = 0.62, n.s. | rrb = 0.75, n.s. | rrb = 0.34, n.s. |
| | Semantic Identification Accuracy - Delayed Recall (%) | 38.9 (25.6) | 79.6 (15.3) | 79.4 (16.5) | rrb = 0.78, p <0.0001 | rrb = 0.78, p <0.0001 | rrb = 0.02, n.s. |
| | Semantic Identification Accuracy - Delayed Recall (z) | -3.1 (2.1) | -0.0 (1.1) | -0.1 (1.2) | rrb = 0.78, p <0.0001 | rrb = 0.76, p <0.0001 | rrb = -0.06, n.s. |
| | Location Error - Immediate Recall (cm) | 3.2 (1.8) | 1.0 (0.4) | 1.9 (1.9) | rrb = -0.82, p <0.0001 | rrb = -0.48, p <0.0001 | rrb = 0.23, n.s. |
| | Location Error - Immediate Recall (z) | 4.8 (5.5) | -0.1 (0.8) | 0.2 (1.3) | rrb = -0.72, p <0.0001 | rrb = -0.67, p <0.0001 | rrb = 0.02, n.s. |
| | Location Error - Delayed Recall (cm) | 5.3 (2.7) | 1.9 (0.9) | 2.3 (1.8) | rrb = -0.72, p <0.0001 | rrb = -0.64, p <0.0001 | rrb = 0.03, n.s. |
| | Location Error - Delayed Recall (z) | 4.0 (3.5) | 0.2 (1.0) | 0.3 (1.5) | rrb = -0.70, p <0.0001 | rrb = -0.68, p <0.0001 | rrb = -0.05, n.s. |
| **ROCF** | Copy Raw Score (%) | 78.5 (30.2) | 98.3 (3.1) | 97.3 (4.9) | rrb = 0.66, p <0.0001 | rrb = 0.57, p <0.0001 | rrb = -0.11, n.s. |
| | Copy Score (z) | -5.5 (8.8) | 0.1 (0.9) | -0.3 (1.6) | rrb = 0.67, p <0.0001 | rrb = 0.57, p <0.0001 | rrb = -0.06, n.s. |
| | Immediate Recall Raw Score (%) | 43.1 (22.8) | 89.9 (11.8) | 86.1 (13.2) | rrb = 0.92, p <0.0001 | rrb = 0.88, p <0.0001 | rrb = -0.17, n.s. |

| | | | | | | | |
|---|---|---|---|---|---|---|---|
| | Immediate Recall Score (z) | -4.4 (2.7) | 0.3 (1.0) | -0.2 (1.3) | rrb = 0.89, p <0.0001 | rrb = 0.83, p <0.0001 | rrb = -0.29, p = 0.0007 |
| **CORSI** | Mean Location Error | 13.0 (5.1) | 6.0 (2.3) | 6.7 (4.7) | rrb = -0.83, p <0.0001 | rrb = -0.72, p <0.0001 | rrb = -0.06, n.s. |
| | Mean Location Error (z) | 2.0 (1.7) | -0.2 (0.8) | 0.3 (1.5) | rrb = -0.77, p <0.0001 | rrb = -0.64, p <0.0001 | rrb = 0.09, n.s. |
| **DSST** | Number of Correct Responses | 14.8 (7.0) | 34.6 (9.3) | 39.5 (12.3) | rrb = 0.89, p <0.0001 | rrb = 0.93, p <0.0001 | rrb = 0.22, n.s. |
| | Number of Correct Responses (z) | -2.3 (1.1) | -0.2 (0.9) | -0.0 (1.2) | rrb = 0.86, p <0.0001 | rrb = 0.87, p <0.0001 | rrb = 0.10, n.s. |
| **TMT** | TMT A Completion Time (s) | 50.1 (26.5) | 29.0 (11.0) | 25.7 (8.2) | rrb = -0.79, p <0.0001 | rrb = -0.86, p <0.0001 | rrb = -0.23, n.s. |
| | TMT A Completion Time (z) | 4.3 (5.3) | 0.5 (1.8) | 0.2 (1.3) | rrb = -0.79, p <0.0001 | rrb = -0.82, p <0.0001 | rrb = -0.12, n.s. |
| | TMT B Completion Time (s) | 155.5 (150.9) | 40.1 (11.4) | 37.6 (13.1) | rrb = -0.88, p <0.0001 | rrb = -0.90, p <0.0001 | rrb = -0.16, n.s. |
| | TMT B Completion Time (z) | 13.8 (21.2) | 0.3 (1.2) | 0.1 (1.2) | rrb = -0.89, p <0.0001 | rrb = -0.89, p <0.0001 | rrb = -0.08, n.s. |
| | Average TMT Completion Time (s) | 102.8 (87.8) | 34.5 (9.9) | 31.6 (9.5) | rrb = -0.89, p <0.0001 | rrb = -0.92, p <0.0001 | rrb = -0.19, n.s. |
| | Average TMT Completion Time (z) | 10.3 (14.6) | 0.4 (1.3) | 0.1 (1.2) | rrb = -0.89, p <0.0001 | rrb = -0.90, p <0.0001 | rrb = -0.10, n.s. |
| | Motor speed (Time to connect two diagnal dots) (s) | 6.5 (4.0) | 3.8 (1.5) | 4.4 (3.4) | rrb = -0.79, p <0.0001 | rrb = -0.57, p <0.0001 | rrb = -0.03, n.s. |
| | Motor speed (Time to connect two diagnal dots) (z) | 2.2 (3.7) | 0.0 (1.4) | 0.3 (1.6) | rrb = -0.78, p <0.0001 | rrb = -0.61, p <0.0001 | rrb = 0.03, n.s. |

#### Supplementary Methods 2 | Processing pipeline for plasma biomarkers

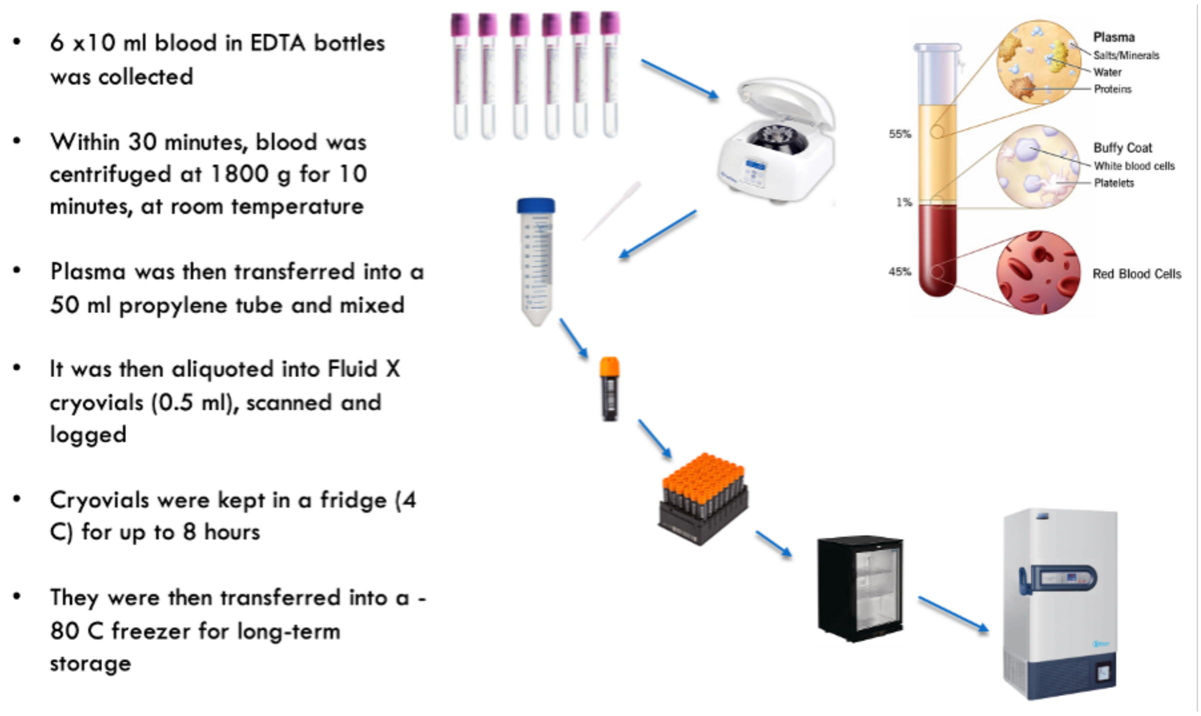

Plasma Aβ40, Aβ42, GFAP, and NfL were measured by Single molecule array (Simoa) technology using the Neurology 4-plexE assay on an HD-X analyser (Quanterix), according to manufacturer's instructions. Plasma pTau181 was also measured by Simoa using the pTau-181 Advantage assay on an HD-X analyser (Quanterix). Briefly, samples were thawed at 21°C, and centrifuged at 10,000 RCF for five minutes at 21°C. On-board the instrument, samples were diluted 1:4 with sample diluent and bound to paramagnetic beads coated with a capture antibody specific for human Aβ40, Aβ42, GFAP, NfL and pTau-181. Aβ40-, Aβ42-, GFAP-, NfL- and pTau181-bound beads were then incubated with a biotinylated anti-Aβ40, anti-Aβ42, anti-GFAP, anti-NfL and anti-pTau181 detection antibodies in turn conjugated to streptavidin-β-galactosidase complex. Subsequent hydrolysis reaction with a resorufin β-D-galactopyranoside substrate produces a fluorescent signal proportional to the concentration of Aβ40, Aβ42, GFAP, NfL and pTau181 present. Singlicate measurements were taken of each sample. Sample concentrations were extrapolated from a standard curve, fitted using a 4-parameter logistic algorithm. Intra-assay and inter-assay coefficients of variation (CVs) were less than 10% and 15% respectively, as determined by 8 quality controls following the same principles.

**Supplementary Figure 1 | Group comparison for all digital cognitive metrics in Oxford Memory Task (OMT) and Object-in-Scene Task (OIS).**

All metrics were normalised based on age-matched normative data.

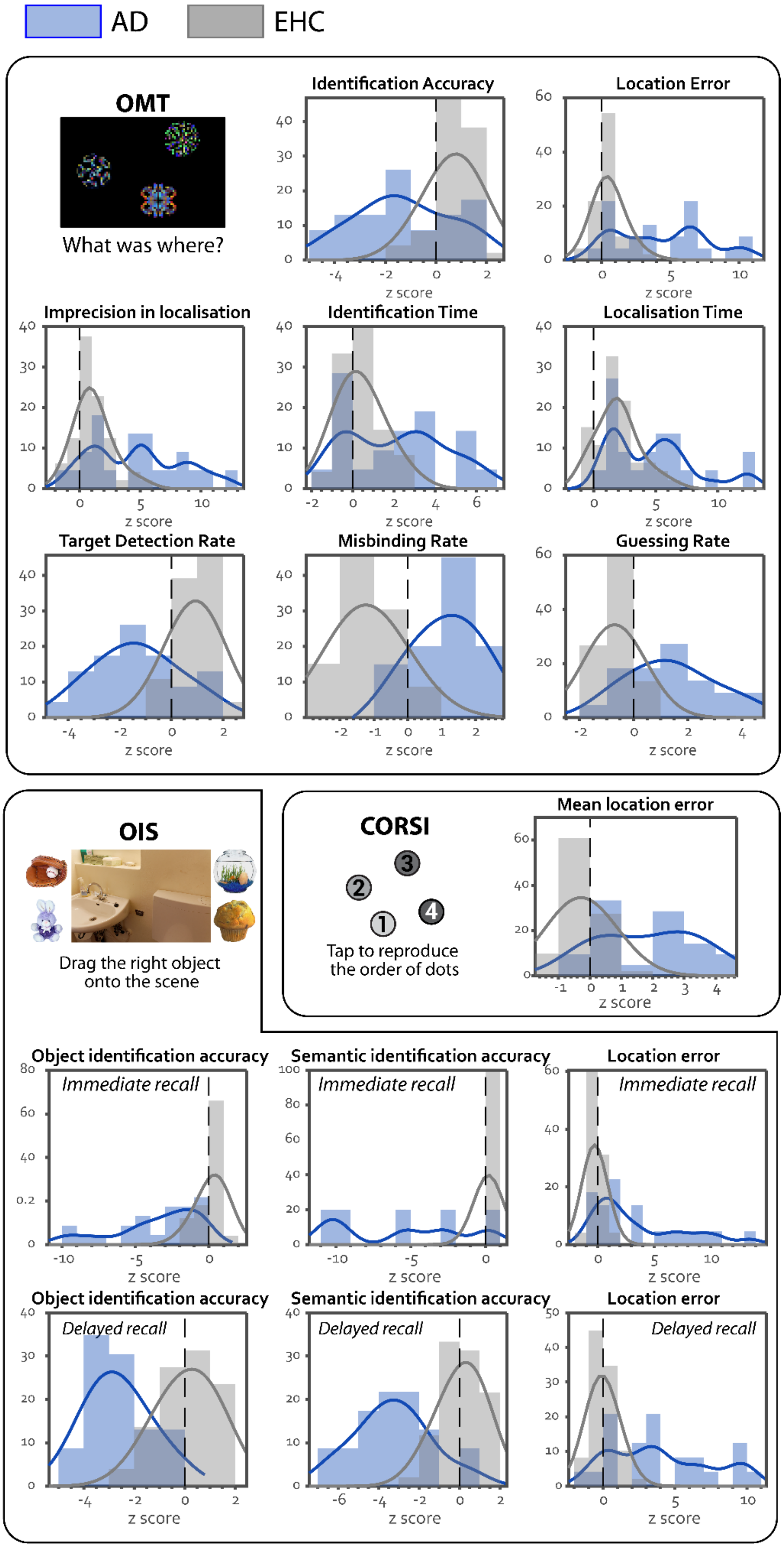

**Supplementary Figure 2 | Group comparison for all digital cognitive metrics in ROCF, DSST and TMT.**

ROCF = Rey–Osterrieth Complex Figure, DSST = Digit Symbol Substitution Task, TMT = Trail Making Test.

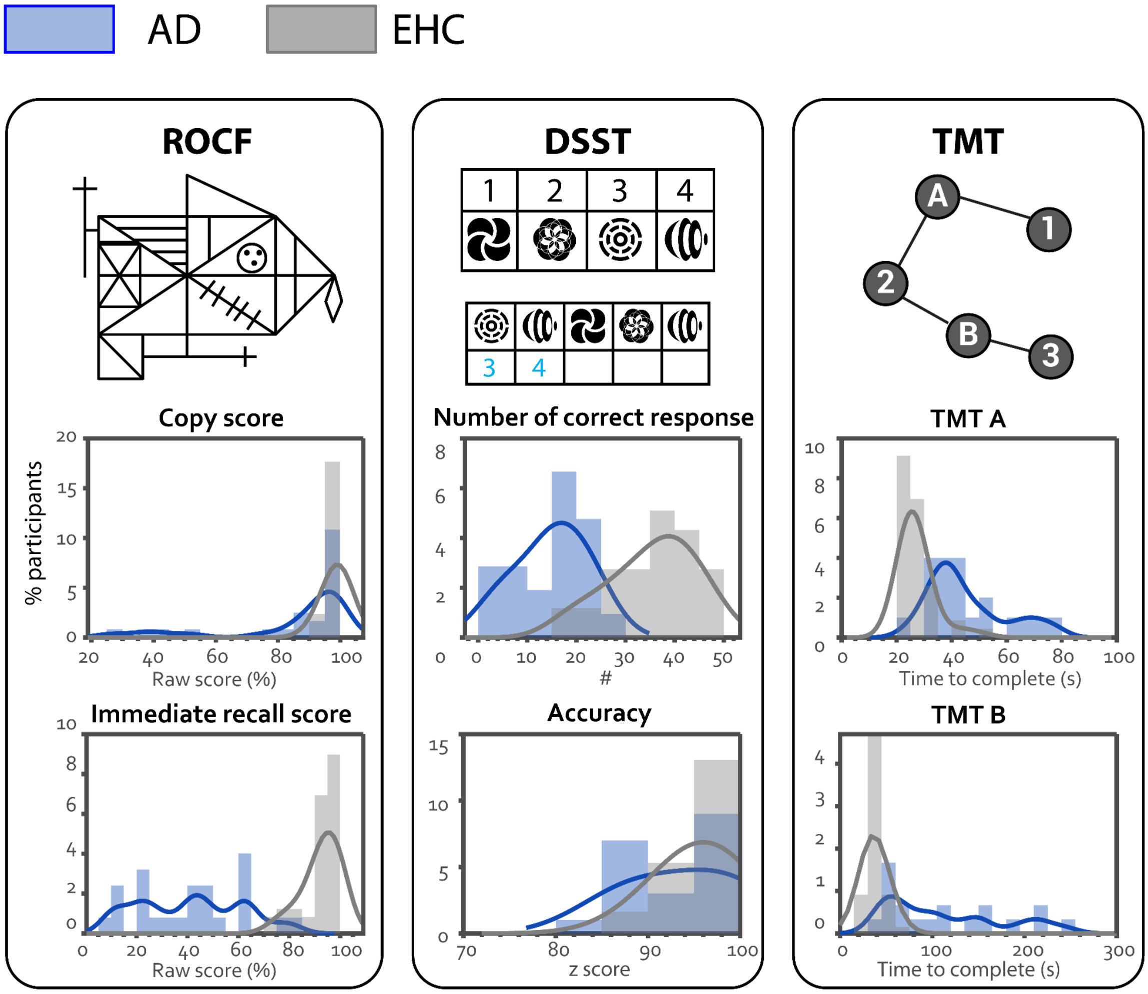

**Supplementary Figure 3 | Apathy and depression are not strongly correlated with plasma biomarkers or performance at digital cognitive tests**

Network plot of relationships between questionnaire-derived apathy (AMI, Apathy Motivation Index), depression (GDS, Geriatric Depression Scale), all plasma biomarkers and online digital cognitive tasks (clustered on the bottom left). Associations are presented in graded colours, where red is associated with a negative correlation and light blue with a positive correlation. The shorter distance between two metrics indicates a stronger relationship (larger correlation coefficient). For online tasks, only one metric was selected per task, according to the highest effect size in discriminating between groups.

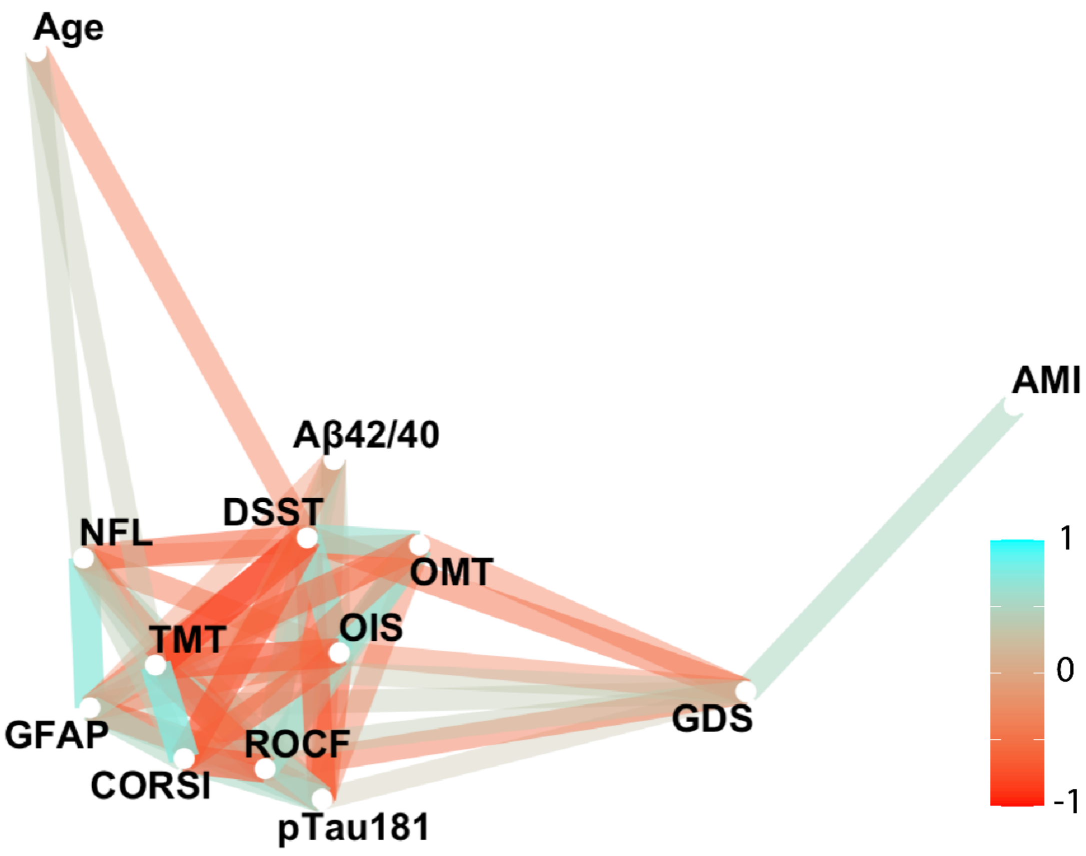

**Supplementary Figure 4 | Which cognitive metric or plasma biomarker best predicts pTau181 or Aβ42/40 levels?**

**(a) Which cognitive metric or plasma biomarker best predicts pTau181? DSST was the winner.** NfL and Aβ42/40 ratio significantly predict the level of pTau181, but ranked behind all digital tasks, with the exception of OMT. **(b) Ranked biomarkers and digital cognitive metrics in predicting Aβ42/40 ratio.** pTau181 was the only metric which significantly predicted Aβ42/40 ratio. OMT = Identification accuracy of the Oxford Memory Task, OIS = Object Identification Accuracy in Immediate Recall of the Object-in-Scene Memory Task, ROCF = recall of the Rey–Osterrieth Complex Figure, DSST = number of correct responses of the Digit Symbol Substitution Task, TMT = average reaction time of the Trail Making Test, CORSI = average location error of the Freestyle Corsi Block Task. **(c) Best combinations of biomarkers and digital cognitive metrics in predicting group (top), pTau181 (middle) and Aβ42/40 ratio (bottom).** AUC = area under the curve. AIC = Akaike information criterion.

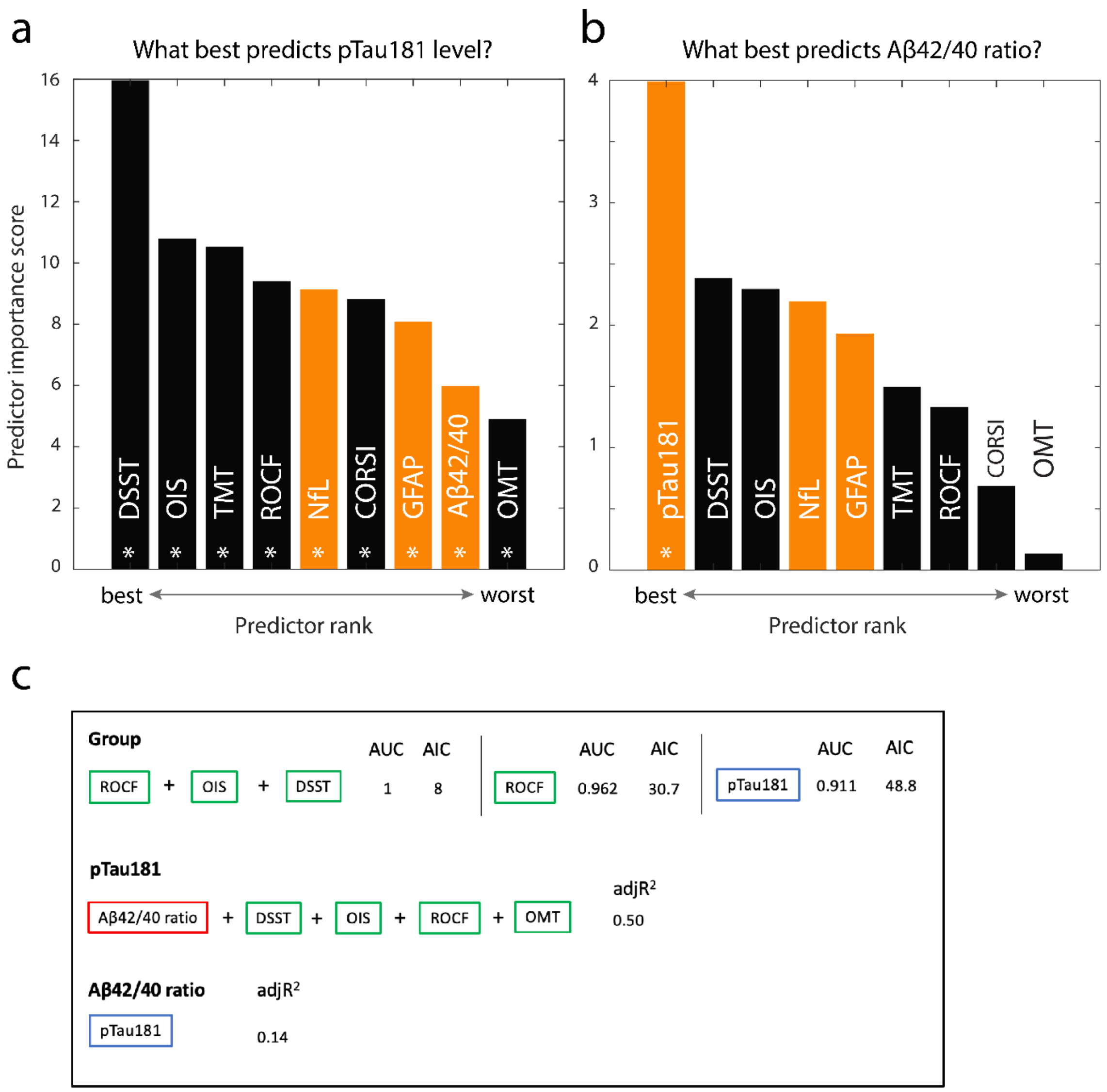

#### References for Supplementary Materials:

[1] Office for National Statistics. Ethnicity facts and figures of UK: Socioeconomic status 2020.
[2] Adler N, Stewart J. The MacArthur scale of subjective social status. MacArthur Research Network on SES & Health., San Francisco: 2007.
[3] Operario D, Adler NE, Williams DR. Subjective social status: reliability and predictive utility for global health. Psychology & Health 2004;19:237–46. https://doi.org/10.1080/08870440310001638098.
[4] Präg P, Mills MC, Wittek R. Subjective socioeconomic status and health in cross-national comparison. Social Science & Medicine 2016;149:84–92. https://doi.org/10.1016/j.socscimed.2015.11.044.
